# Heterogeneity of acetylcholine receptor autoantibody-mediated complement activity in patients with myasthenia gravis

**DOI:** 10.1101/2021.10.05.21264566

**Authors:** Abeer H. Obaid, Chryssa Zografou, Douangsone D. Vadysirisack, Bailey Munro-Sheldon, Miriam L. Fichtner, Bhaskar Roy, William M. Philbrick, Jeffrey L. Bennett, Richard J. Nowak, Kevin C. O’Connor

**Affiliations:** Department of Neurology, Yale School of Medicine, New Haven, CT 06511 USA; Department of Immunobiology, Yale School of Medicine, New Haven, CT 06511 USA; Institute of Biomedical Studies, Baylor University, Waco, TX 76706 USA; UCB Pharma, Cambridge, MA 02140, USA; Departments of Neurology and Ophthalmology, Programs in Neuroscience and Immunology, University of Colorado Anschutz Medical Campus, Aurora, Colorado, 80045, USA

**Keywords:** [132] Autoimmune diseases, [179] Myasthenia, Complement, Autoantibodies

## Abstract

**Background:** Autoantibodies targeting the acetylcholine receptor (AChR), found in patients with myasthenia gravis (MG), mediate pathology through three mechanisms: complement-directed tissue damage, blocking of the acetylcholine binding site, and internalization of the AChR. Clinical assays, used to diagnose and monitor patients, measure only autoantibody binding. Consequently, they are limited in providing association with disease burden, understanding of mechanistic heterogeneity, and monitoring therapeutic response.

**Objective:** Develop a cell-based assay that measures AChR autoantibody-mediated complement membrane attack complex (MAC) formation.

**Methods:** An HEK293T cell line—modified using CRISPR/Cas9 genome editing to disrupt expression of the complement regulator genes (CD46, CD55 and CD59)—was used to measure AChR autoantibody-mediated MAC formation via flow cytometry.

**Results:** Serum samples (n=155) from 96 clinically confirmed AChR MG patients, representing a wide range of disease burden and autoantibody titer, were tested along with 32 healthy donor (HD) samples. AChR autoantibodies were detected in 139 of the 155 (89.7%) MG samples via a cell-based assay. Of the 139 AChR positive samples, autoantibody-mediated MAC formation was detected in 83 (59.7%), while MAC formation was undetectable in the HD group or AChR positive samples with low autoantibody levels. MAC formation was positively associated with autoantibody binding in most patient samples; ratios (MFI) of MAC formation to AChR autoantibody binding ranged between 0.27–48, with a median of 0.79 and interquartile range of 0.43 (0.58–1.1). However, the distribution of ratios was asymmetric and included extreme values; 16 samples were beyond the 10–90 percentile, with high-MAC to low-AChR autoantibody binding ratio or the reverse. Correlation between MAC formation and clinical disease scores suggested a modest positive association (*rho*=0.34, p=0.0023), which included a subset of outliers that did not follow this pattern. MAC formation did not associate with exposure to immunotherapy, thymectomy, or MG subtypes defined by age-of-onset.

**Conclusions:** A novel assay for evaluating AChR autoantibody-mediated complement activity was developed. A subset of patients that lack association between MAC formation and autoantibody binding or disease burden was identified. The assay may provide a better understanding of the heterogeneous autoantibody molecular pathology and identify patients expected to benefit from complement inhibitor therapy.

## Introduction

The impairment in autoimmune myasthenia gravis (MG) is caused by autoantibodies that target components of the neuromuscular junction (NMJ)^1-3^. The most common MG subtype is characterized by autoantibodies targeting the acetylcholine receptor (AChR)^1^. Detection of AChR autoantibodies confirms a diagnosis of MG, however titers (serum concentration) vary widely among patients and within individuals during their disease, and it is generally recognized that cross-sectional measurements of titer do not correlate well with disease severity among different patients^4-7^.

AChR autoantibodies are broadly polyclonal and heterogeneous in their specificity; MG serum can include autoantibodies that can recognize any of the four different AChR subunits (α, β, ε/γ, or δ)^8^. AChR autoantibodies can use three distinct mechanisms^9-15^ to effect pathology: (i) complement-directed tissue damage, (ii) blocking the binding site for acetylcholine on the receptor, and (iii) modulation (internalization) of the AChR. Furthermore, there may also be a subgroup of binding-only antibodies that recognize AChR in clinical assays but do not have any pathogenic properties.

In addition to the lack of understanding concerning the disconnect between autoantibody titer and disease severity, gaps in our knowledge have recently become more consequential because new therapeutics specifically targeting the pathogenic mechanisms of AChR autoantibodies—including complement inhibitors—have been introduced^16^. Eculizumab is a recently approved biological therapeutic for the treatment of generalized AChR MG^17^; it inhibits autoantibody-mediated activity of the terminal complement cascade. The phase III clinical trial showed efficacy, but approximately 40% of AChR autoantibody-positive patients did not meet the trial endpoint without longer-term treatment, and some experienced exacerbation events requiring rescue therapy^17-19^. These data are challenging to reconcile as the poor responders had measurable circulating AChR autoantibodies. This trial also highlights the limitations of AChR autoantibody titer as a biomarker and importantly underscores the need for further understanding of AChR autoantibody-mediated pathogenic mechanisms so that response to treatments can be better predicted. To that end, we sought to develop an *in vitro* assay that measures AChR autoantibody-mediated complement activation.

## Methods

### Standard Protocol Approvals, Registrations, and Patient Consent

This study was approved by Yale University’s Institutional Review Board (clinicaltrials.gov || NCT03792659). Informed written consent was received from all participating patients prior to inclusion in this study. Serum was collected from AChR MG patients at the Yale Medicine Myasthenia Gravis Clinic^20^. Demographics and clinical data are presented in **Table e1**.

### CRISPR/Cas9 genome editing of HEK293T cells

CD46, CD55, and CD59 CRISPR sgRNAs (**Table e2**) were cloned into the pSpCas9(BB) -2A-Puro (PX459) V2.0^21^ vector (a gift from Dr. Feng Zhang (Addgene plasmid # 62988)) and were transfected into human embryonic kidney (HEK)293T cells (ATCC CRL3216). The cells were harvested after 48 hours, and dilutions were used to derive single cell clonal cell lines. These clones were then tested phenotypically for the deletions of CD46, CD55, and CD59 via PCR, Sanger sequencing, and flow cytometry. Clones that exhibited deletions underwent FACS sorting to ensure the selection of cells with the three deletions.

### Autoantibody binding live cell-based assay (CBA)

Wild type (WT), triple KO HEK293T, or Chinese hamster ovary (CHO) cells (CCL-61) were transfected with adult AChR (2α, β, δ, and ε) and rapsyn-GFP plasmids (generous gifts from Drs. Angela Vincent, David Beeson, and Patrick Waters) using branched polyethylenimine or Lipofectamine 2000 (CHO only). After a 24-hour incubation, the media was refreshed, and the CBA was conducted the following day. Cells were exposed to serum (1:20 dilution) from healthy donors (HD) or MG patients or monoclonal antibodies (mAbs) (10 µg/ml). Bound antibodies were detected with an anti-human IgG Fcγ (309-605-008; Jackson ImmunoResearch) with a LSR Fortessa (BD Biosciences). FlowJo software (v10.6.2) was used for analysis. In serial dilution testing, some serum samples can provide aberrantly low CBA Δ mean florescence intensity (MFI) values when tested at 1:20, due to assay saturation and/or interference. Accordingly, these data points were not considered in the analyses. For testing AQP4 binding, cells were transiently transfected with an AQP4-GFP (M1 isoform) plasmid^22^, using the CBA conditions described above.

### Complement cell-based assay

Complement competent normal human serum (NHS) and factor B-depleted human serum were both purchased from Complement Technologies (Tyler, TX). The same lot was used within experiments. Human recombinant mAbs were expressed as we have previously described^23^.

CD46, CD55, CD59 KO HEK293T cells or CHO cells were prepared as outlined in the live CBA. Cells were washed twice and resuspended in complete DMEM. Cells were exposed to AChR MG patient serum, HD serum (both heat-inactivated), or control mAbs suspended in 25% NHS or Factor B depleted serum. MAC formation was allowed to proceed for three hours at 37ºC, then fixed with 2% PFA at 4ºC for 15 minutes. The cells were then stained with anti-C9 neoantigen (Hycult, Cat# HM2264), followed by washing and secondary staining (APC (Cat # 407109) or PE IgG2a clone RMG2a-62 (Cat# 407109)). FACS analysis was conducted as described for the binding CBA.

### Statistical analysis

Threshold values for the CBA and complement CBA positivity were determined by the application of a cutoff value, determined by calculating the mean +4 SD of HD subject samples. For descriptive statistics, mean, median, or percentage were used as appropriate. Comparison between two groups was performed using the Mann-Whitney U test, and the Spearman correlation was used to examine the relationship between two parameters. A box-whisker plot (whisker representing 10–90 percentile) was used to graphically represent the distribution of the ratios to examine the relationship between MAC and AChR autoantibody binding. P-values below 0.05 were considered significant. All statistical analyses were performed on Graphpad Prism software (v9.1.2). STROBE cohort reporting guidelines were used.^24^

### Data Availability

Anonymized data will be shared upon request from qualified investigators and completion of materials transfer agreements.

## Results

### Measuring autoantibody-mediated MAC formation using a CBA platform

AChR autoantibodies can be measured with live CBAs^25^. In this assay format, recombinant human AChR subunits and the clustering protein, rapsyn, are expressed in human embryonic kidney (HEK) cells and tested for binding of autoantibodies. We sought to measure AChR autoantibody-mediated complement activity by modifying the clustered AChR CBA. To that end, the assay was performed by using autoantibodies (recombinant mAbs or autoimmune patient-derived serum, in which the complement was heat-inactivated) and normal human serum (NHS) as a consistent complement source.

We first evaluated antibody-dependent MAC formation, by detecting a C9 neo-epitope, with the modified CBA using an established positive control; specifically, human aquaporin (AQP4) autoantibodies from patients with neuromyelitis optica spectrum disorder (NMOSD), which have been demonstrated to efficiently fix complement *in vitro*^26^. AQP4 was expressed in HEK293T cells, then a human AQP4-specific recombinant IgG1 subclass mAb (mAb-58^27^) or serum from an AQP4 autoantibody positive NMOSD patient was added along with NHS. MAC formation was detected when the AQP4-specific mAb or NMOSD serum and NHS were present. MAC formation was not detected in the absence of a AQP4-specific antibody source, absence of NHS, or when heat-inactivated NHS was included (**eFigure 1A, eFigure 1B**). We next tested AChR autoantibody-mediated MAC formation under the same conditions, but with AChR-rapsyn expressed in HEK293T cells. An AChR-specific human recombinant IgG1 subclass mAb (mAb-637^28^) and serum from an AChR autoantibody positive patient (MG-87) were tested. In contrast to the results obtained in the AQP4 system, no AChR-autoantibody-dependent MAC formation was detected (**eFigure 1C**).

Most human cells express the complement regulators CD46, CD55, and CD59, which can interfere with complement deposition. Therefore, we tested the complement CBA with CHO cells, which do not express these regulators^29^. An AChR-specific cell-based binding assay with CHO cells was established, but nonspecific background binding by human IgG prevented its use for complement detection (**eFigure 2A-D**). A previous study demonstrated that the human HAP1 cell line, in which the expression of complement regulators was disrupted through gene editing, provided increased complement component deposition^30^. Thus, we reasoned that a genome-edited HEK293T cell line, in which the complement regulators CD46, CD55, and CD59 were not expressed, would be an effective tool in evaluating AChR autoantibody-mediated complement activity. Accordingly, we produced a modified HEK293T cell line, using CRISPR/Cas9 genome editing to target these genes (**eFigure 3A-C, eTable 2**). We then confirmed that autoantibody binding was uncompromised in the modified HEK293T cells (**eFigure 4A**) and that AChR was consistently expressed (**eFigure 4B**).

### AChR autoantibody-mediated MAC formation in the absence of complement regulators

The CD46/55/59 KO HEK293T engineered cells were then tested in the complement CBA, first using the AQP4 antigen. The AQP4-specific mAb-58 or serum from an AQP4 autoantibody positive NMOSD patient, was added to the cells and tested for MAC formation as described for the WT HEK cells. MAC formation was detected when the AQP4-specific mAb or serum was added together with NHS. Conversely, MAC formation was not detected in the absence of either the AQP4 autoantibody source or NHS, or when the mAb or serum was added with heat-inactivated NHS (**Figure 1A**). The AChR-specific mAb-637, serum from a patient with MG, and healthy controls, which were tested under the same conditions, did not initiate MAC formation in the AQP4 complement CBA (**Figure 1A**).

**Figure 1.**
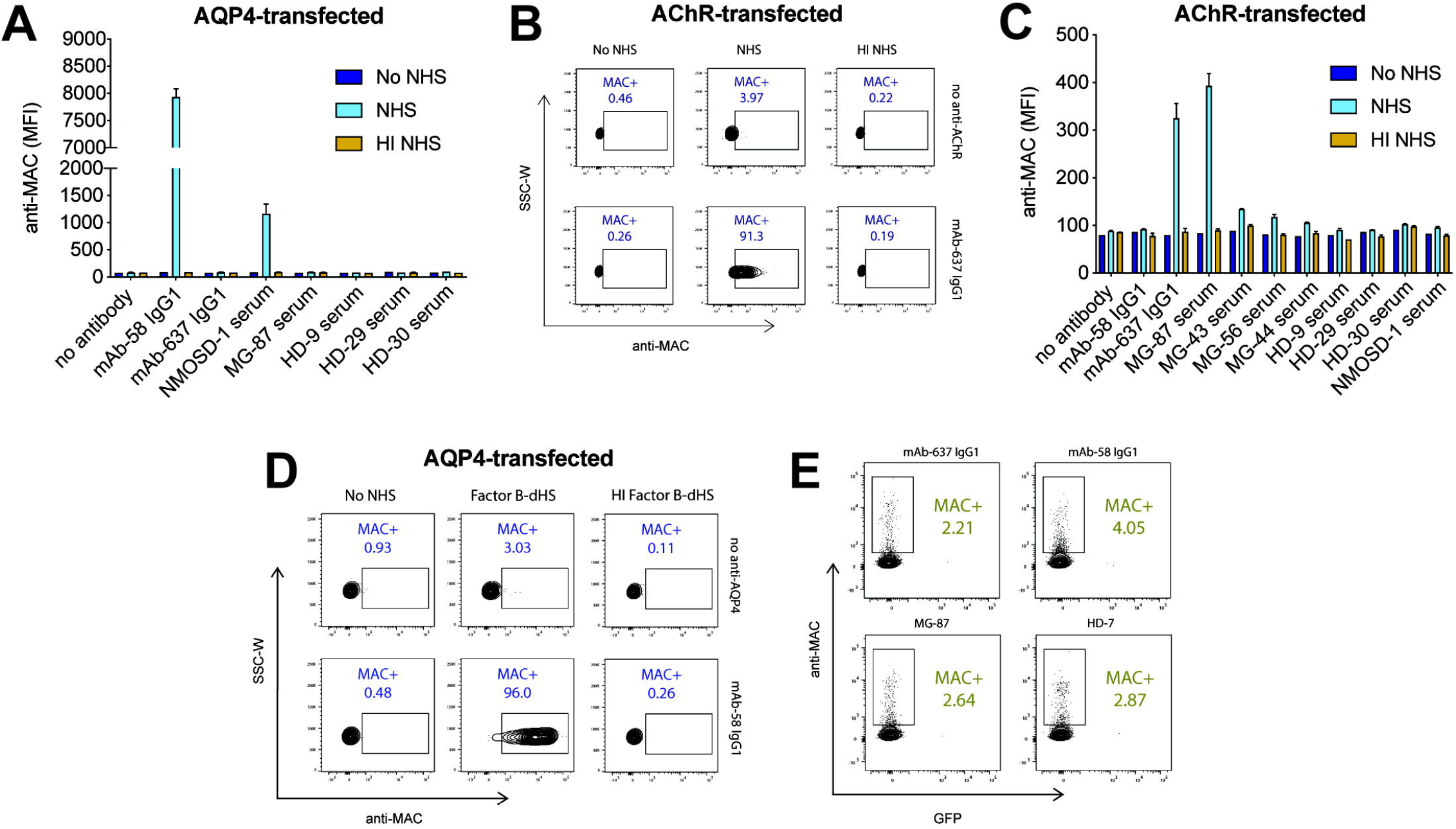
Autoantibody-mediated complement fixation assay with CD46/55/59 KO HEK293T cells. Autoantibodies were tested for their ability to mediate complement activation through measuring MAC formation via FACS on live cells, transfected with self-antigens, in which the complement regulators (CD46, CD55, and CD59) were knocked out. Normal human serum (NHS) was added as consistent complement source. Control conditions included omitted NHS or heat-inactivated NHS. All tested patient or HD serum samples were heat-inactivated. **A**. Measurement of MAC formation using an *in vitro* complement CBA with AQP4 transfected CD46/55/59 KO HEK293T cells. Complement fixation was tested with AQP4-specific autoantibody sources (anti-AQP4 monoclonal mAb-58 IgG1 and NMOSD serum) and negative control autoantibody sources (anti-AChR mAb-637 IgG1 and serum from MG patients and HDs). NHS and HI NHS bars represents the mean of duplicate experimental conditions while No NHS bars represent singlets. **B**. Representative FACS analysis of AChR transfected CD46/55/59 KO HEK293T cells using an *in vitro* complement CBA. AChR transfected cells were tested without (top row) or with the anti-AChR mAb-637 IgG1 (bottom row). **C**. Measurement of MAC formation using the complement CBA with AChR transfected CD46/55/59 KO HEK293T. Complement fixation was tested with AChR-specific autoantibody sources (anti-AChR mAb-637 IgG1 and serum from MG patients) and negative control autoantibody sources (anti-AQP4 monoclonal mAb-58 IgG1 and serum from NMOSD patients and HDs). NHS and HI NHS bars represents the mean of duplicate experimental conditions while No NHS bars represent singlets. **D**. Factor B depleted NHS was used in the complement CBA to test whether the alternative pathway contributed to autoantibody-mediated MAC formation. AQP4 transfected cells were tested without (top row) or with the anti-AQP4 monoclonal mAb-58 IgG1 (bottom row). **E**. Non-transfected CD46/55/59 KO HEK293T cells were tested in the complement CBA using antigen-specific monoclonal antibodies (mAb-637 or mAb-58) or serum from MG patients and HDs to test for the requirement of autoantigens in MAC formation. Samples used in panels **A–E:** mAb-58 IgG1 (anti-AQP4 monoclonal), NMOSD patient (NMOSD-1), mAb-637 IgG1 (anti-AChR monoclonal), MG patient serum (MG-87, MG-43, MG-56, MG-44), and healthy donors (HD-9, HD-29, HD-30). Blank—no serum, NHS—normal human serum, HI NHS—heat-inactivated normal human serum, Factor B-dHS—factor B depleted human serum, HI Factor B-dHS—heat-inactivated factor B depleted human serum.

To test the complement CBA with the modified cells for AChR autoantibody-mediated MAC formation, AChR and rapsyn were co-expressed, and then the 637-mAb was added to the cells and tested for MAC formation. MAC formation was detected when the AChR-specific mAb and NHS were present (**Figure 1B**). We next tested MAC formation with serum samples from four AChR autoantibody positive MG patients and four non-MG serum samples. Two were derived from MG patients (**eTable 1**) with AChR autoantibody titers—as measured by the clinical diagnostic radioimmunoassay (RIA)—that were exceptionally high (MG-87 and MG-56, 43nM and 88nM respectively) and two had mid-low range titers (MG-43 and MG-44, 14.3nM and 1.47nM respectively). Serum from MG-87 mediated robust MAC formation (**Figure 1C**). Serum from MG-56 and MG-43 mediated weak MAC formation and MG-44 showed none. HD serum, NMOSD serum, and the AQP4-specific mAb-58 did not mediate MAC formation on the AChR-expressing cells (**Figure 1C**).

To confirm that the antibody-mediated classical complement pathway was generating the MAC, we tested the contribution of the alternative pathway. This was achieved through inhibiting the alternative pathway by using NHS, in which factor B—required for the alternative pathway—was depleted. The MAC was detected when AQP4-specific mAb-58 and factor B depleted serum were present but not in the absence of either the autoantibody or factor B depleted serum, or when the mAb was added with heat-inactivated factor B depleted serum (**Figure 1D**). To confirm that the self-antigen was driving autoantibody binding and subsequent MAC formation, non-transfected cells were tested in the complement CBA using the AQP4-specific mAb-58, AChR-specific mAb-637, HD serum, and an MG patient serum sample (MG-87). MAC formation was below or at background levels using the non-transfected cells, indicating the requirement of antigen recognition in initiating antibody-mediated MAC formation (**Figure 1E**).

### Testing MG patient serum for autoantibody-mediated MAC formation

Having demonstrated that the complement CBA could measure AChR autoantibody-mediated classical pathway MAC formation, we next tested MG serum samples to explore how AChR autoantibody-mediated complement activity was represented in a heterogeneous population of MG patients. MAC formation was tested using heat inactivated serum from 30 HD subjects (n=32 samples) and from 96 patients (n=155 samples) with a confirmed diagnosis of AChR MG (**eTable 1**). While all the MG patients were diagnosed with the AChR disease subtype, we specifically included patient samples that were AChR autoantibody positive and others that were negative at the time of collection (**eTable 1**). An assay positivity cutoff was established using the MAC MFI from the HD cohort (156± 14 MFI); samples with an MFI > mean + 4SD of the HD cohort were considered positive. Serum from the HD cohort did not show any measurable AChR autoantibody-mediated MAC formation (**Figure 2A**). A subset (83/155, 53.5%) of the MG patient-derived samples, demonstrated AChR autoantibody-mediated MAC formation (**Figure 2A**). To determine whether those samples, which did not mediate MAC formation, included autoantibodies that were detectable by CBA, we next evaluated the association between MAC formation and AChR autoantibody binding by comparing the complement CBA results to those of the antibody-binding CBA. There was a significant correlation (*rho*=0.897; p<0.0001) observed between MAC formation and AChR autoantibody binding (**Figure 2B**). Samples for which there was no detectable binding in the CBA did not mediate detectable MAC formation in the complement CBA (MAC formation was detected in 83/139 (59.7%) AChR positive samples). Additionally, samples that provided a positive, yet low, signal in the binding CBA did not mediate detectable MAC formation. Samples that were collected longitudinally presented a similar association pattern (**eFigure 5**).

**Figure 2.**
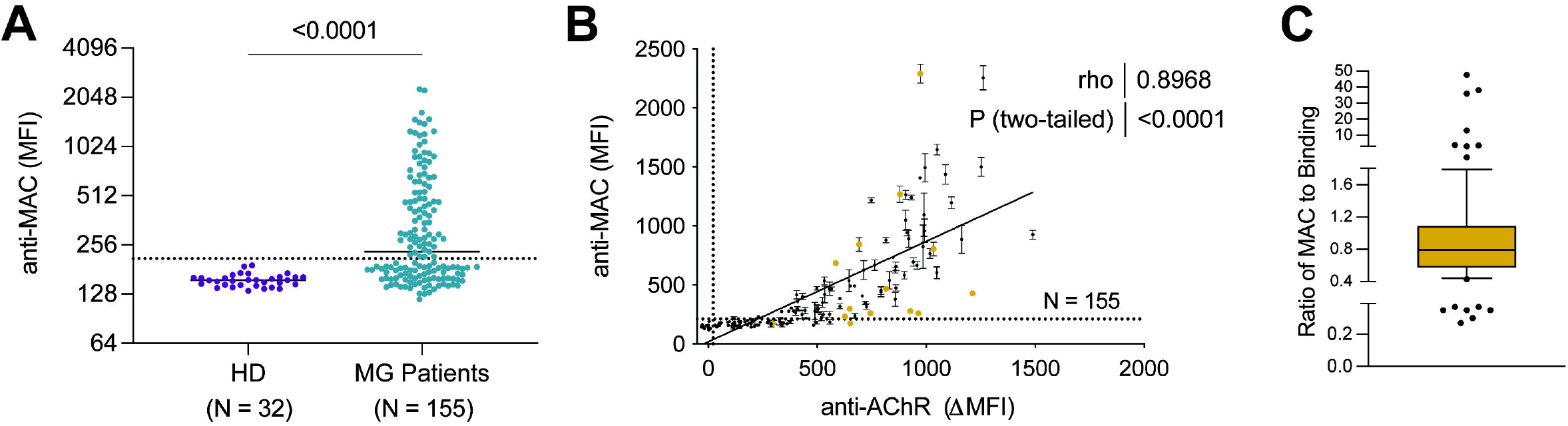
AChR autoantibody-mediated complement formation in MG patients. AChR autoantibodies in serum were tested for their ability to mediate complement activation through measuring MAC formation on live AChR-expressing CD46/55/59 KO HEK293T cells. **A**. Comparison of MAC formation using serum samples (n=32) from HDs and serum samples (n=155) from AChR MG patients. The cutoff for positivity was set at the mean MFI + 4 STD (210.9 MFI) of the HD samples. Each data point represents the mean of triplicate experimental conditions. **B**. Correlation between MAC formation and AChR autoantibody binding by CBA. Spearman correlation was used to calculate the relationship. The binding CBA cutoff for positivity was set at the mean MFI +4STD (21.91 ΔMFI) of the HD samples. Each data point represents the mean of triplicate experimental conditions. Colored (gold) points represent samples selected for further evaluation (*see* **Figure 3**). **C**. Graphical representation of heterogeneity in MAC formation and AChR autoantibody binding of MG samples. Cross sectional samples (n=89) were analyzed through a ratio of MAC formation and autoantibody binding, to observe disassociation between AChR autoantibody binding and complement formation. The box-whisker plot shows the median (0.79) and 10–90 percentile (outlined by the whiskers) of the MAC to AChR autoantibody binding ratios. The dots represent data outside of the 10–90 percentile. The Y-axis is not to scale and was divided in three sections for better visual representation. All samples were measured in triplicate, and those with negative ΔMFI binding values (n=5) were excluded from the analysis.

Despite a strong correlation between MAC formation and AChR autoantibody binding (**Figure 2B**), we noted some samples with high MAC values despite relatively low AChR autoantibody binding, and some with high AChR autoantibody binding with relatively low MAC values, which suggests a degree of heterogeneity among this population. To further explore the disassociation between MAC formation and AChR autoantibody binding, we examined the distribution of the ratio of MAC formation to AChR autoantibody binding. The ratios ranged between 0.27–48, with a median of 0.79 and interquartile range of 0.43 (0.58–1.1), with an asymmetric, non-normal distribution with extreme values (skewness 5.1 and kurtosis 26 (leptokurtic)). The box-whisker plot (**Figure 2C**) shows relatively higher or lower ratios between MAC and AChR autoantibody binding beyond 10–90 percentiles, graphically representing the aforementioned heterogeneity among this population. These collective results suggest that, for the majority of MG patient serum AChR autoantibodies, CBA binding positively associates with autoantibody-mediated CBA MAC formation, but a subset did not follow this pattern.

### Comparing titers of MG patient serum autoantibody binding and autoantibody-mediated MAC formation

While CBA-based binding and MAC formation were closely associated for many samples, the results also suggest that a subset of patients may harbor AChR binding autoantibodies that range broadly in their ability to mediate MAC formation. Accordingly, 14 samples were selected for further study (**Figure 2B**), based on their observed association/disassociation between the MAC formation and AChR autoantibody binding. Each sample was tested for AChR autoantibody binding and MAC formation at various serum dilutions (**Figure 3, eFigure 6**). The binding CBA, which tested serum over three-fold dilutions from 1:20–1:1350, showed a range of binding titers. The complement CBA, which tested serum over two-fold dilutions from 1:20–1:320, showed a wide range of MAC formation. Of interest was that several samples showed similarly strong AChR autoantibody binding profiles but differed considerably in their MAC formation capability. For example, samples MG-20 and MG-34 have comparable binding profiles but considerably differ in their MAC formation capability; MG-20 mediated robust MAC formation at three of the serum dilutions tested, while MG-34 did not provide a signal above the cutoff at any serum dilution. Similarly, MG-49, MG-51, and MG-89 provided strong binding signals, but only MG-89 provided a strong MAC formation signal. These collective results imply that the autoantibodies harbored by these patients have differing pathogenic capacities irrespective of binding capability.

**Figure 3.**
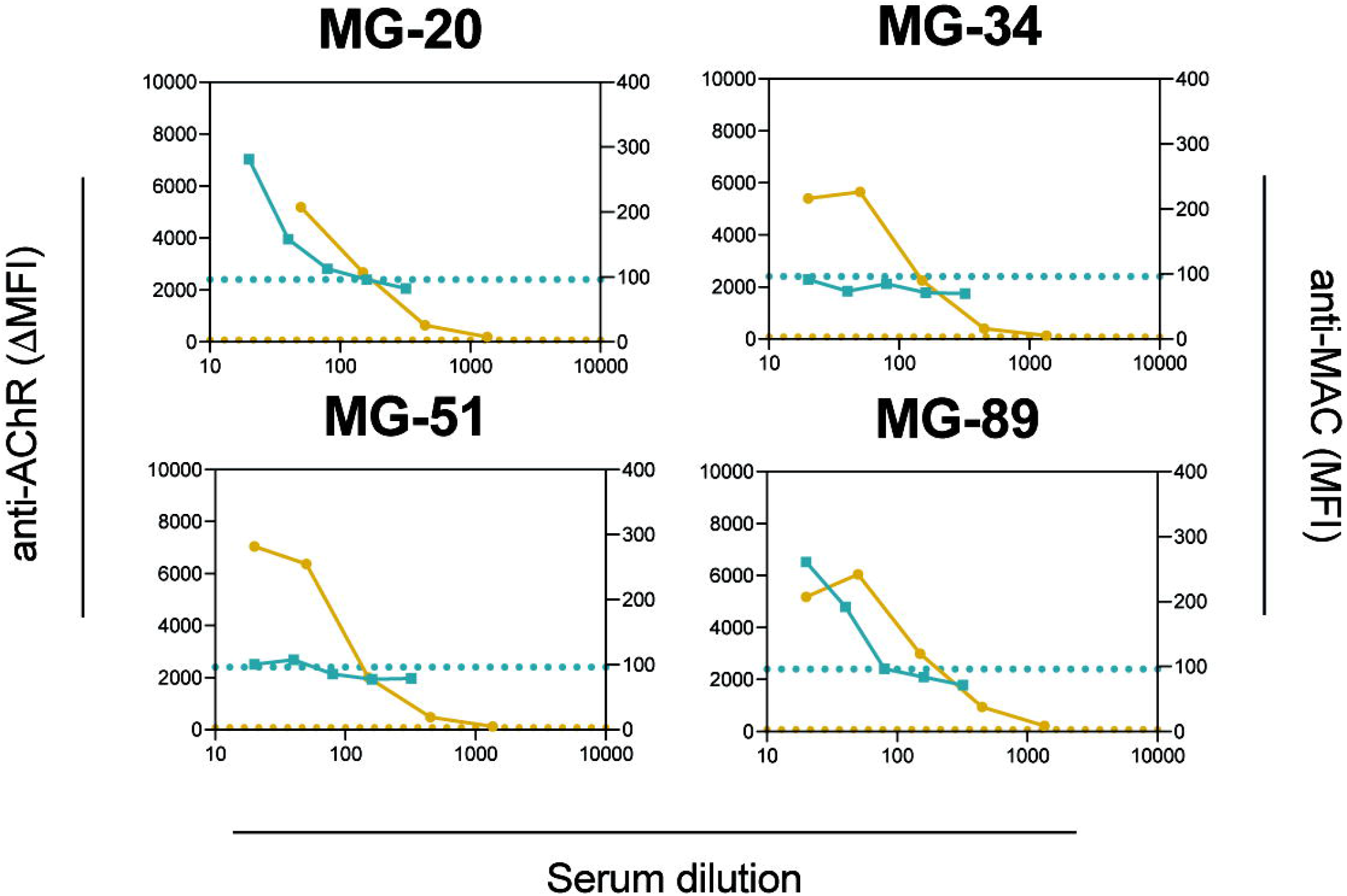
Heterogeneous AChR autoantibody-mediated complement formation. Selected samples (indicated by golden points in **Figure 2B**) were examined side-by-side for both MAC formation and CBA binding. Each graph shows the data collected from individual serum samples. The X-axis represents the serum dilution of the sample tested for complement CBA (MAC) formation (blue dots) or CBA binding (golden dots). For AChR autoantibody binding (left y-axis), samples were tested at serum dilutions of 1:20 plus 4 additional 3-fold dilutions (1:50, 1:150, 1:450, and 1:1350). For MAC formation (right y-axis), samples were tested at 2-fold serial dilutions (1:20, 1:40, 1:80, 1:160, and 1:320). Each data point represents the mean of experimental triplicate experimental conditions. Dotted horizonal lines mark the positive reactivity cutoff for complement (blue) and CBA binding (golden). Cutoffs were calculated using the mean MFI + 4STD of the HD samples (complement and CBA 96.01 MFI and 75.35 ΔMFI respectively). Data from additional samples are shown in **eFigure 6**.

### Association between AChR autoantibody assay data and disease severity, treatment, or MG subtypes

We first sought to explore the relationship between disease severity and autoantibody binding and autoantibody-mediated MAC formation. We first categorized the MG patients into two subgroups using the MGFA clinical classification: asymptomatic/ocular disease (MGFA = 0/I) and mild-to-severe generalized disease (MGFA ≥ II) severity (**eTable 1**). Patients with higher disease severity showed both higher CBA binding and MAC formation when compared to those with minimal disease burden as measured by the MGFA clinical classification (p <0.0001 and p <0.0001 respectively) (**Figure 4A and B**). We next assessed the relationship between AChR autoantibody binding and MAC formation with the MG composite score (MGC)—a weighted disease severity score scale—by Spearman correlation. Both binding and MAC formation showed mild-to-moderate correlation with MGC (*rho* = 0.437, p = 0.0001; *rho* = 0.3512, p = 0.0023 respectively) (**Figure 4C and 4D**). Samples that were collected longitudinally presented a similar qualitative association between MAC formation, binding, and disease severity (**eFigure 5**). However, several samples in both the cross sectional (**Figure 4**) and longitudinal (**eFigure 5**) analyses stood out because of their lack of a positive association, reflecting a similar disassociation to that which we observed between binding and MAC formation. For example, MG–37 had a moderate to high disease score, yet low antibody binding and MAC formation values. The opposite case was also found (MG-52) where both binding and MAC formation values were high relative to other samples, but the disease scores were low (**eFigure 5**). We also examined if there were differences in autoantibody binding and autoantibody-complement mediated MAC formation in specific MG patient subsets. Differences between treatment naïve and patients receiving immunotherapy, early-onset MG (EOMG) and late-onset MG (LOMG), and patients with or without thymectomy (**eFigure 7**) were evaluated. None of the subsets showed any significant differences in terms of MAC formation and CBA binding. Collectively, these findings suggest that antibody-mediated complement activity moderately associates with disease burden, but some patients with mild and others with more severe disease have respectively high or low autoantibody-mediated MAC formation activity.

**Figure 4.**
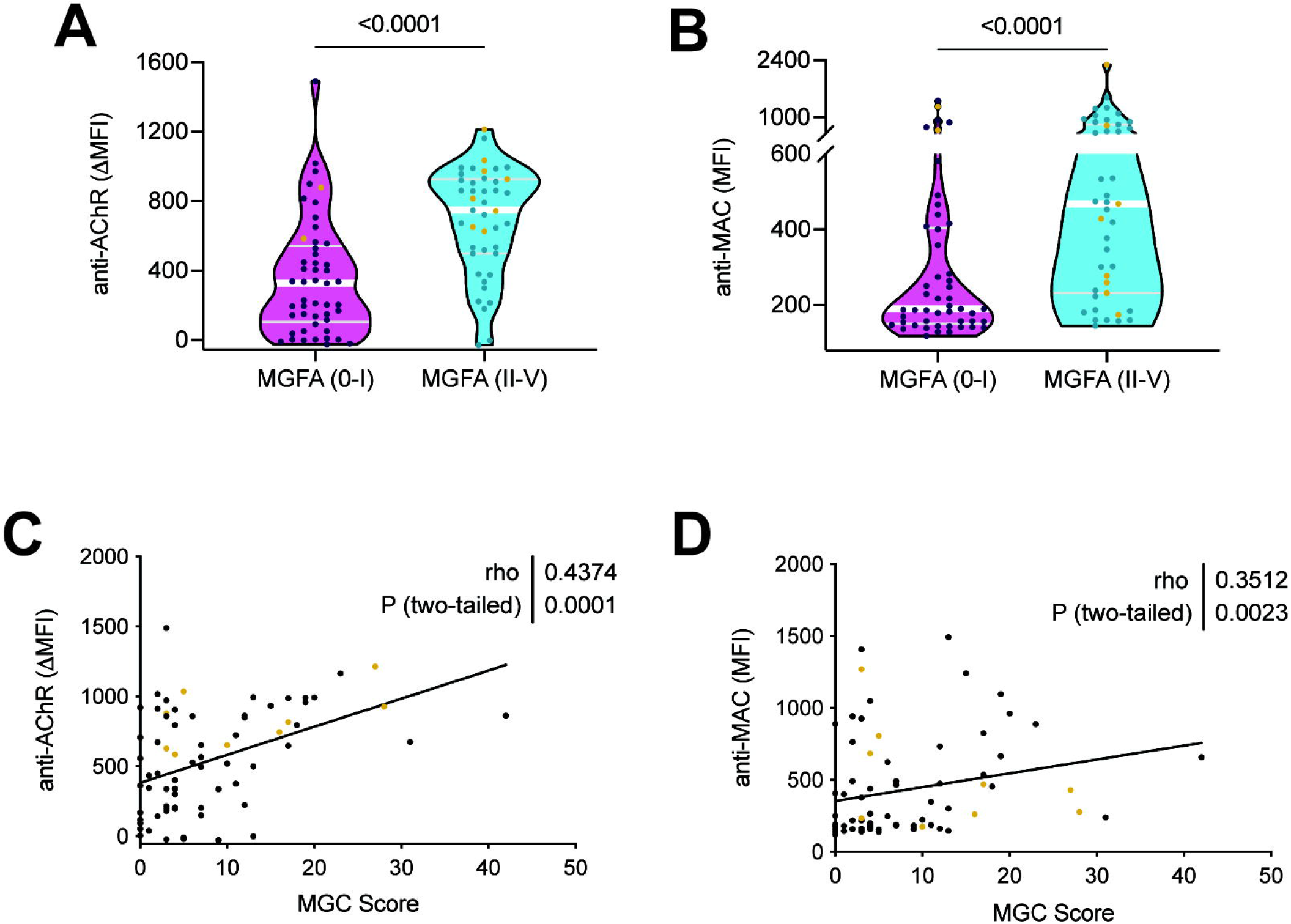
Correlation between clinical disease measurements and autoantibody-mediated complement formation. Cross-sectional samples were assessed for correlation between MGFA classification or MG composite (MGC) score and the binding CBA or complement CBA data. **A–B**. Samples with low disease severity (MGFA 0/I) were compared to samples with higher disease severity (MGFA II-V) for differences in **(A)** autoantibody binding (median MGFA (0/I): 327.3 (II-V): 748.2; p-value <0.0001) or **(B)** MAC formation (median MGFA (0/I): 190.3 (II-V): 468.3; p-value <0.0001). **C–D**. Spearman correlation was used to calculate the relationship between cross-sectional sample MGC scores and **(C)** autoantibody binding (*rho* 0.437, p-value = 0.0001) or **(D)** MAC formation (*rho* 0.351, p-value = 0.0023). Each data point in **A–D** represents the mean of triplicate experimental conditions. White and grey lines in **A** and **B** represent the median and quartiles respectively.

## Discussion

Detection of AChR autoantibodies confirms a diagnosis of MG, however the discordance between titer and disease severity may be, in part, due to how AChR autoantibody titer is measured. Specifically, the clinical assays used to diagnose patients and provide titer values are unable to discriminate between the detection of the autoantibodies and their pathogenic properties (effector functions). Importantly, nonpathogenic AChR binding-only autoantibodies, which would be detected in clinical assays, may also exist, further complicating the disconnect between disease severity and autoantibody titer. Such nonpathogenic binding autoantibodies have been identified in other autoimmune conditions including pemphigus^31^ and NMOSD^32^. The nonpathogenic NMOSD autoantibodies, which are thought to be poor at activating complement due to altered Fc regions, compete with pathogenic autoantibodies for antigen binding^32^. Our data revealed a positive association between autoantibody binding and MAC formation, suggesting that a considerable fraction of circulating AChR autoantibodies can initiate the complement cascade. However, binding alone did not account for the variation observed in complement activity. We reason that heterogeneous autoantibody characteristics, among other factors, may play an integral role in determining MAC formation efficiency. Such characteristics would minimally include subunit and epitope specificity, affinity, IgG subclass usage, Fc structure, and post-translation modifications (PTM), all of which can alter complement activating capability in addition to structure and stability^33, 34^. Although it is currently unclear how AChR autoantibody heterogeneity affects MAC formation in MG patients, parallels to the mechanisms governing AQP4 autoantibody-mediated pathology in NMOSD provide insight. AQP4-binding autoantibodies display broad efficiency in inducing complement-mediated cell death^26^. Here, a combination of distinct properties, including particular epitope binding and the assembly of multimeric antigen platforms, are required for ideal activation of autoantibody complement-mediated cell death^26^. Specifically, AQP4 autoantibodies that target a distinct epitope on the AQP4 extracellular loop C demonstrate remarkably enhanced complement activity when compared to autoantibodies recognizing other epitopes. Furthermore, AQP4 can form supramolecular orthogonal arrays that arrange these epitopes in a manner that benefits autoantibody multimeric complexes, which support autoantibody Fc-Fc interactions that are critical for efficient C1q binding and subsequent complement activity^26^. Whether or not similar mechanisms occur in the context of AChR-specific MG has yet to be explored. However, given the variable complement activity shown here and that AChR is tightly clustered by the intracellular scaffolding protein, rapsyn, such organized formations of self-antigen may support Fc-Fc interactions facilitating efficient complement activation, by AChR autoantibodies recognizing particular—but not yet identified—epitopes. Thus, we propose that circulating AChR autoantibodies include subtypes that contribute to heterogeneous pathogenetic mechanisms and show variable efficiency in affecting these mechanisms, and finally that their composition varies among patients and perhaps longitudinally within a patient. We further suggest that AChR autoantibody subunit specificity and epitope specificity is linked to these effector functions, including activation of the classical complement pathway.

We recognize that this study includes limitations. First, measuring complement formation on live cells with flow cytometry presents challenges given that the cells may be actively lysing or dying and therefore not detected. We empirically arrived at a single timepoint after the addition of NHS that provided measurable differences in MAC formation in the AQP4 and AChR complement CBAs when comparing patient samples and controls. This approach was chosen in consideration of assay ease-of-use. Longitudinal measurements after the addition of NHS or different approaches that measure cumulative cell death, such as Cr^52^ release, may provide additional information on AChR autoantibody-mediated activity, but were outside the scope of our study. Second, autoantibody-mediated MAC formation was not detected in a number of samples, most of which showed lower, but still positive, CBA binding. It is reasonable to conclude that these samples do indeed include AChR autoantibodies capable of mediating MAC formation. However, the sensitivity of the complement CBA may require further optimization to detect their activity. Finally, we recognize that the *in vitro* autoantibody-mediated complement activity may not fully reflect an *in vivo* complement response. We are using an *in vitro* system with diminished expression of complement inhibitory proteins. Our system reflects the capability of the autoantibody to initiate complement but does not consider the varying levels of complement inhibitory protein expression on various muscle cell surfaces. In this regard, the assay only simulates more complex activity at the NMJ; however, the clinical binding CBA also lacks any representation of the NMJ other than antigen expression, but it is recognized as being highly reliable for diagnosis of MG. Further emphasizing this point, it is challenging to understand why it was required to remove the complement regulators, as they are present in the NMJ. It may be related to *in vitro* assay sensitivity, but further study is required to understand this observation. Interestingly, this requirement has been consistently observed in other studies that investigated pathophysiologic properties of AChR autoantibodies^35^ using *in vitro* assays and *in vivo* models; specifically, CD55 knockout mice, which are more susceptible to the effects of pathogenic MG autoantibodies^36-38^. These regulators have also been implicated in ocular MG, where reduced expression levels of CD55 and CD59 in extraocular muscles offer a potential explanation for why these muscle subgroups are highly associated with MG^36^. To reflect the NMJ-specific conditions more accurately, our assay could be complemented by other approaches. For example, motor neurons—the target cells in patients with multifocal motor neuropathy (MMN)—have been used for investigating the pathogenicity of MMN autoantibodies^39^. In similar fashion, functional consequences of complement damage could be assessed with NMJ-specific cell types.

We observed a modest correlation between autoantibody-mediated complement activity and disease burden. While additional mechanisms must be considered, this suggests that in some patients’ complement activity may make major contributions to pathology. Conversely, patients who did not fit this pattern, for example those with high disease burden and low complement activity, may harbor autoantibodies that mediate pathology through other mechanisms (blocking and/or modulating). Furthermore, we did not observe differences in autoantibody-mediated complement activity between EOMG and LOMG, which suggests that the properties of the autoantibodies may be similar in these major MG subsets. Immunomodulatory therapy and thymectomy did not reveal diminished complement activity. Autoantibody titer often persists following such treatments^40, 41^, thus our findings suggest that these treatments may not alter either the properties or relative frequency of complement activating autoantibodies.

It is anticipated that measuring AChR autoantibody-mediated pathogenesis will have clinical utility. The last decade has seen a considerable number of new and existing therapeutics become available for the treatment of MG^16^. Included are biologics that directly target the autoantibody effector function (eculizumab and others)^46^. Clinical trials of these treatments have provided outcomes demonstrating benefit in some patients but unexpected failure in others, thus intensifying challenges to the field, including the need to target the correct therapy to patients from the growing number of options, and the need to inform treatment decisions through understanding the immunopathology. Therapeutic complement inhibition provides one such example of our limited understanding of AChR MG immunomechanisms: it is unclear why eculizumab (approved for the treatment of generalized AChR MG^17^) does not yield clinical benefit in some AChR autoantibody-positive patients^17, 18^, even with longer-term treatment^19^. The AChR autoantibody titers of the patients who responded to eculizumab treatment neither predicted treatment efficacy nor correlated with its progression. In addition to emphasizing the limitations of AChR autoantibody titer as a biomarker, these studies also underscore the need to further understand AChR autoantibody mechanisms so that response to treatments can be better predicted. Future experiments, which include testing samples from complement inhibitor trials, could be performed to investigate whether the complement CBA is valuable in forecasting response. To this end, we suggest a suite of clinical assays that measure the composition of AChR autoantibodies with variable pathogenic mechanisms will be valuable. These assays would ideally measure binding-only, classical complement activation as shown here, and modulating and blocking functions, which have been demonstrated with similar CBA platforms^48, 49^. These collective measurements may be useful for assessing disease progression and serve as an improved MG biomarker when compared to autoantibody binding. Thus, patients would be provided with the opportunity for a more individualized treatment plan that targets their unique autoantibody-mediated pathogenic pathways.

## Data Availability

All data referred to in the manuscript will be made available to qualified investigators upon request, and upon completion of appropriate institutional MTA documentation.

## Acknowledgements

The authors thank Drs. Soumya Yandamuri, Gianvito Masi, Philip Coish, and Ms. Sarah Ohashi for critical reading of the manuscript.

## Notes

**Study funding:** KCO was supported by the National Institute of Allergy and Infectious Diseases of the NIH under award numbers R01-AI114780 and R21 AI164590; through an MGNet pilot grant awarded through the Rare Diseases Clinical Research Consortia of the NIH (award number U54-NS115054) and by a High Impact Clinical Research and Scientific Pilot Project award from the Myasthenia Gravis Foundation of American (MGFA). This project was also supported, in part, by grants to KCO and RJN, from Ra Pharmaceuticals, now a part of UCB. JLB was supported by the National Eye Institute R01-EY022936 and R21-032399. MLF was supported in part by the SPIN award through Grifols and has further been supported by a DFG Research fellowship (FI 2471/1-1).

### Competing Interest Statement

Abeer H. Obaid reports no disclosures.
Dr. Chryssa Zografou reports no disclosures.
Dr. Douangsone D. Vadysirisack is an employee of UCB Ra Pharma.
Dr. Bailey Munro-Sheldon reports no disclosures.
Dr. Miriam L. Fichtner reports no disclosures.
Dr. Roy is a consultant for Alexion Pharmaceuticals, (now part of AstraZeneca), and Takeda Pharmaceuticals.
Dr. William M. Philbrick reports no disclosures.
Dr. Jeffrey L. Bennett reports payment consultation fees for study design/consultation from Viela Bio, now part of Horizon Therapeutics; and personal fees from Mitsubishi Tanabe Pharma Corporation; Reistone Bio, AbbVie, Alexion; Chugai, Clene Nanomedicine, Genentech, Genzyme, Mitsubishi-Tanabe, and Roche; grants from Novartis, Mallinckrodt; and has a patent for Aquaporumab issued.
Dr. Richard J. Nowak has received research support from the National Institutes of Health (NIH), Genentech, Alexion Pharmaceuticals, argenx, Annexon Biosciences, UCB Ra Pharmaceuticals, Myasthenia Gravis Foundation of America, Momenta, Immunovant, Grifols, and Viela Bio, now part of Horizon Therapeutics. RJN has served as consultant/advisor for Alexion Pharmaceuticals, argenx, Cabaletta Bio, CSL Behring, Grifols, Ra Pharmaceuticals, now a part of UCB Pharma, Immunovant, Momenta, and Viela Bio, now part of Horizon Therapeutics.
Dr. Kevin C. OConnor has received research support from Ra Pharma, now part of UCB Pharma, and Alexion, now part of AstraZeneca and Viela Bio, now part of Horizon therapeutics. KCO is a consultant and equity shareholder of Cabaletta Bio, and is the recipient of a sponsored research subaward from the University of Pennsylvania, the primary financial sponsor of which is Cabaletta Bio. KCO has served as consultant/advisor for Alexion Pharmaceuticals, now part of AstraZeneca, and for Roche, and he has received speaking fees from Alexion, Roche, Genentech, and Viela Bio, now part of Horizon Therapeutics.

### Funding Statement

KCO was supported by the National Institute of Allergy and Infectious Diseases of the NIH under award numbers R01-AI114780 and R21 AI164590; through an MGNet pilot grant awarded through the Rare Diseases Clinical Research Consortia of the NIH (award number U54-NS115054) and by a High Impact Clinical Research and Scientific Pilot Project award from the Myasthenia Gravis Foundation of American (MGFA). This project was supported, in part, by grants to KCO and RJN, from Ra Pharmaceuticals, now a part of UCB. JLB was supported by the National Eye Institute R01-EY022936 and R21-032399. MLF was supported in part by the SPIN award through Grifols and has further been supported by a DFG Research fellowship (FI 2471/1-1)

### Author Declarations

This study was approved by Yale University's Institutional Review Board (clinicaltrials.gov || NCT03792659). Informed written consent was received from all participating patients prior to inclusion in this study.

### Summary of Updates

New clinical analyses New author (Dr. Bhaskar Roy)

