## Supplemental Data for "Heterogeneity of acetylcholine receptor autoantibody-mediated complement activity in patients with myasthenia gravis"

**eFigure 1.** Autoantibody-mediated complement fixation cell-based assay (CBA) using HEK293T cells.

**eFigure 2.** Representative flow cytometry plots of cell-based assay experiments with CHO cells.

**eFigure 3.** Design and testing of HEK293T cells engineered to lack surface expression of CD46, CD55 and CD59.

**eFigure 4.** Functional testing of the CD46/55/59 KO HEK cells in the CBA.

**eFigure 5.** Longitudinal changes in AChR autoantibody binding and MAC formation.

**eFigure 6.** Heterogeneous AChR autoantibody-mediated complement formation.

**eFigure 7**. CBA binding and MAC formation comparison between MG subgroups.

**eTable 1.** Demographics and clinical data of patient and control groups.

**eTable 2.** CRISPR gRNA sequences for the generation of CD46/55/59 KO HEK293T.

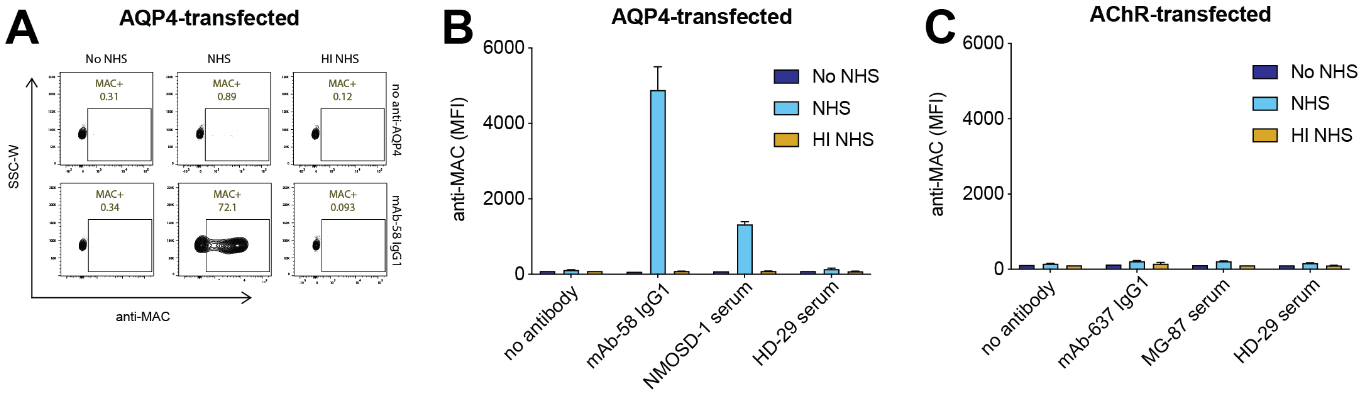

**eFigure 1. Autoantibody-mediated complement fixation cell-based assay (CBA) using HEK293T cells.** Autoantibodies were tested for their ability to mediate complement activation by measuring MAC formation on live cells (HEK293T cells) transfected with AQP4 or AChR. **A.** Representative FACS plots showing MAC formation in AQP4 transfected cells using an *in vitro* complement CBA. Very low MAC assemblies were detected in the absence of a complement source (normal human serum (NHS)), or heat inactivated normal human serum (HI NHS), while MAC-positive cells were observed when a monoclonal antibody, (anti-AQP4 monoclonal (mAb-58 IgG1; 10 µg/mL)), was added with normal human serum (*NHS is included as a consistent and controlled source of human complement*). *Gating strategy:* HEK293T → single cell (SSC-W vs SSC-H) → single cells (FSC-W vs FSC-H) → antigen (*measured by GFP*) positive cells. Values indicate the fraction (%) of cells in the gated population (box). **B and C.** Measurement of MAC deposition using an *in vitro* complement CBA on AQP4 or AChR transfected cells. Cells were incubated with both the antibody and NHS for 3 hours at 37°C. Complement fixation was activated by the addition of NHS and anti-AChR or anti-AQP4 antibody sources (anti-AQP4 monoclonal (mAb-58 IgG1; 10 µg/mL), NMO patient serum (NMOSD-1; 1:20 dilution), anti-AChR monoclonal (mAb-637 IgG1; 10µg/mL), and MG patient serum (MG-87; 1:20 dilution). Controls included no addition of antibody source or healthy donor serum (HD-29; 1:20 dilution). Very low MAC assemblies were detected with AChR transfected cells. NHS and HI NHS bars represents the mean of duplicate experimental conditions while No NHS bars represent singlets. Blank- no serum, NHS- normal human serum, HI NHS- heat inactivated normal human serum.

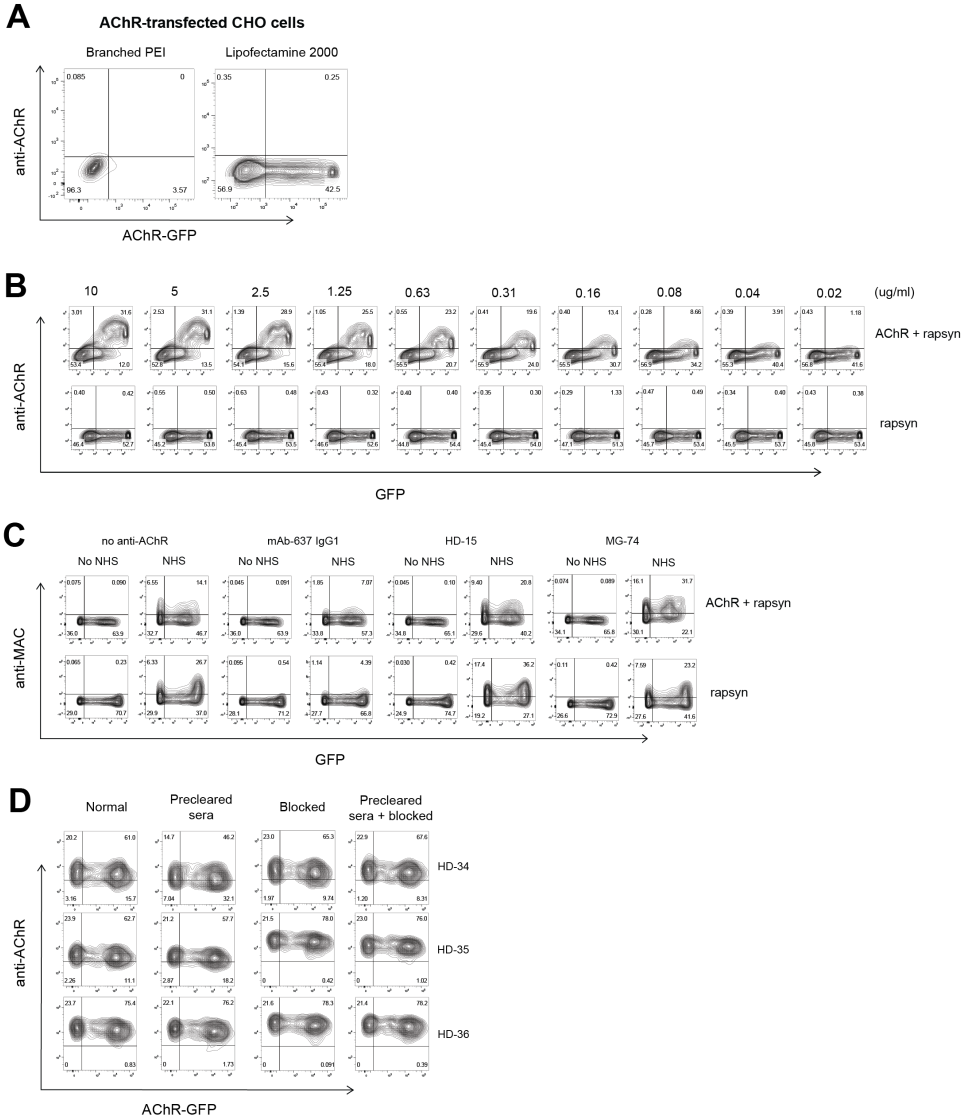

**eFigure 2: Representative flow cytometry plots of cell-based assay experiments with CHO cells.** Testing of an AChR-specific cell-based binding and complement assay with CHO cells **A.** Transfection of the AChR (co-transfected with Rapsyn) of CHO cells was tested with PEI and Lipofectamine 2000. CHO cells can be transfected with AChR using PEI but the efficiency was low; the efficiency was increased by approximately 20-fold using lipofectamine. **B.** Assessment of binding capacity of either AChR or rapsyn-only transfected CHO cells with the anti-AChR monoclonal (mAb-637 IgG1) over a broad range of concentrations (10 - 0.02 µg/ml). The AChR-specific mAb-637 could be detected on AChR-transfected CHO cells over a broad range of concentrations, while rapsyn-only transfected cells showed no binding signal, collectively indicated that a binding CBA could be performed with CHO cells. **C.** Whether CHO cells could be used for the complement (MAC) deposition assay was then investigated. AChR-transfected CHO cells were tested with the complement CBA protocol. NHS was used as complement source and either a HI HD serum, a HI MG serum or mAb-637 IgG1 were used as the autoantibody source. The legend indicates the conditions used for each plot. Anti-MAC was used for detection. High non-specific background binding was observed with the NHS on both the AChR and rapsyn-only transfected cells. This was likely due to non-specific binding of human IgG to the CHO cells from the NHS and/or the heat-inactivated sera. **D.** To test reduction of nonspecific binding, three HD samples were tested on rapsyn-only transfected CHO cells. The sera were pre-cleared by incubation with the CHO cells or CHO cells were blocked with goat serum, or both conditions were tested. Neither blocking nor preclearing nor a combination of both had any effect on reducing the nonspecific background binding. Values in each quadrant in **A-D** indicate the fraction (%) of cells in the population. Anti-human IgG was used for detection in **A, B** and **D**. Anti-MAC (anti-C9 neoantigen) was used in **C**.

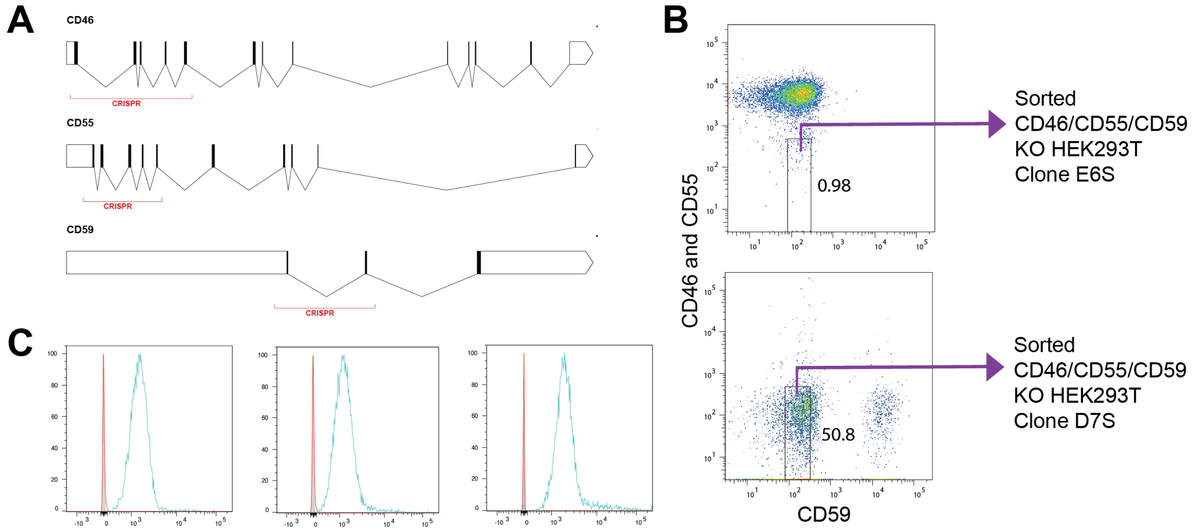

**eFigure 3. Design and testing of HEK293T cells engineered to lack surface expression of CD46, CD55 and CD59. A.** Schematic of sgRNA primer positions for CRISPR/Cas9. Several single cell clones underwent multiple rounds of CRISPR editing and were then sorted for triple negative populations**. B.** Representative FACS analysis of two single cell derived CD46/CD55/CD59 CRISPR/Cas9 genome edited HEK293T clones which were sorted for CD46/55/59 deficient cells. *Gating strategy:* CD46/55/59 KO HEK293T → single cell (SSC-W vs SSC-H) → single cells (FSC-W vs FSC-H) → CD59/CD55/CD46 negative population sorted. Of these clones, protein expression on the cell surface was assessed via FACS analysis. Values indicate the fraction (%) of cells in the gated population (box). **C**. FACS analysis of CD46, CD55 and CD59 expression on WT (blue) and CD46/55/59 KO HEK293T cell (red). No expression of the genes was observed on the KO line as indicated by its peak overlapping with the unstained and secondary only control (grey). Primary antibodies (Biolegend) for flow cytometry analysis included CD46 clone TRA-2-10 (Cat# 352403), CD55 clone JS11 (Cat# 311301), CD59 clone H19 (Cat# 304701) and secondary antibodies PE Cy7 IgG1 clone RMG1-1 (Cat# 406613) and APC (Cat # 407109) or PE IgG2a clone RMG2a-62 (Cat# 407109). The HEK293T is a hypotriploid human cell line that exhibits a 64 modal chromosome number. This complexity created a significant challenge for the generation of the complement regulator knockout compared to other cells lines such as HAP1 which are haploid. Consequently, we frequently monitored the absence of surface expression of CD46, CD55 or CD59 and continued to use the cell line at early passages (<10) to avoid the development of a heterogeneous culture. Further genetic engineering and analysis will be required to generate a stable single cell-derived clone.

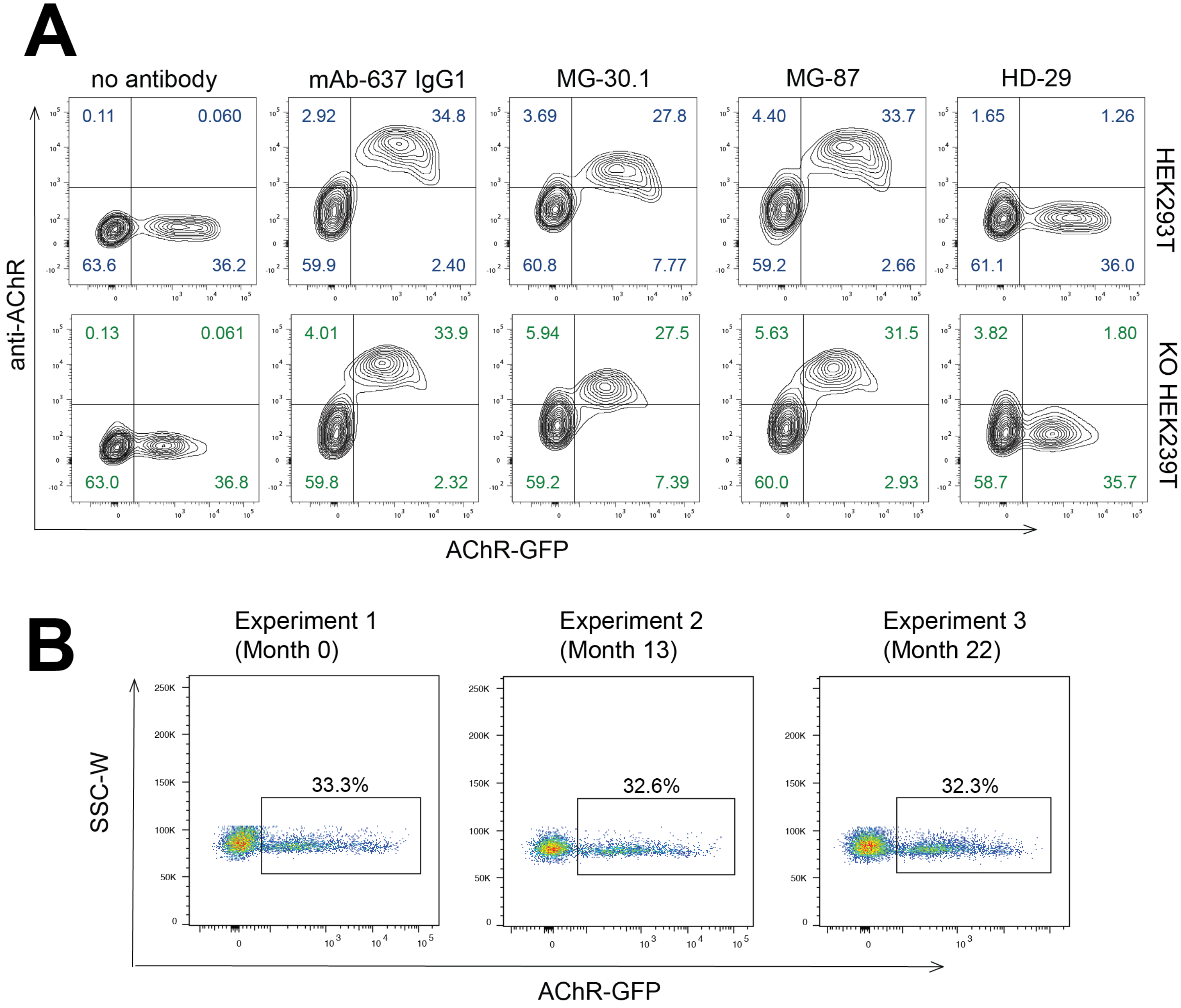

**eFigure 4: Functional testing of the CD46/55/59 KO HEK cells in the CBA.** Testing was performed to compare the CD46/55/59 KO HEK and WT cells in the binding CBA and evaluate consistency of AChR expression longitudinally. First, to confirm that AChR binding by the autoantibodies was not compromised in the modified HEK293T cells, the CD46/55/59 KO cells were compared with WT HEK239T cells in a live CBA. **A.** Both cell types (HEK293T or CD46/55/59 KO HEK293T cells) were transiently transfected with AChR and were tested with no antibody, AChR-specific human recombinant mAb-637 IgG1 (10ug/ml), MG patient serum (diluted 1:20), or HD serum (diluted 1:20). Binding was observed when cells were exposed to antibodies or patient serum. Values refer to the fraction (%) of cells in each quadrant. WT HEK and triple KO cells showed indistinguishable antibody binding patterns with mAb-637 and MG serum. No autoantibody binding with HD serum was observed in either cell line. Anti-human IgG was used for detection of AChR autoantibodies. To confirm that the CD46/55/59 KO HEK293T cells performed consistently over time, AChR expression was measured in independent experiments performed over several months. **B.** AChR transfection and expression was tested through measuring the fraction of KO HEK293T cells expressing rapsyn-GFP, as a proxy for AChR, over a 22-month period. Representative flow cytometry plots of AChR transfection across multiple experiments are shown. AChR transfection and expression was consistent across different experiments conducted 13- and 22-months after experiment 1 (month 0).

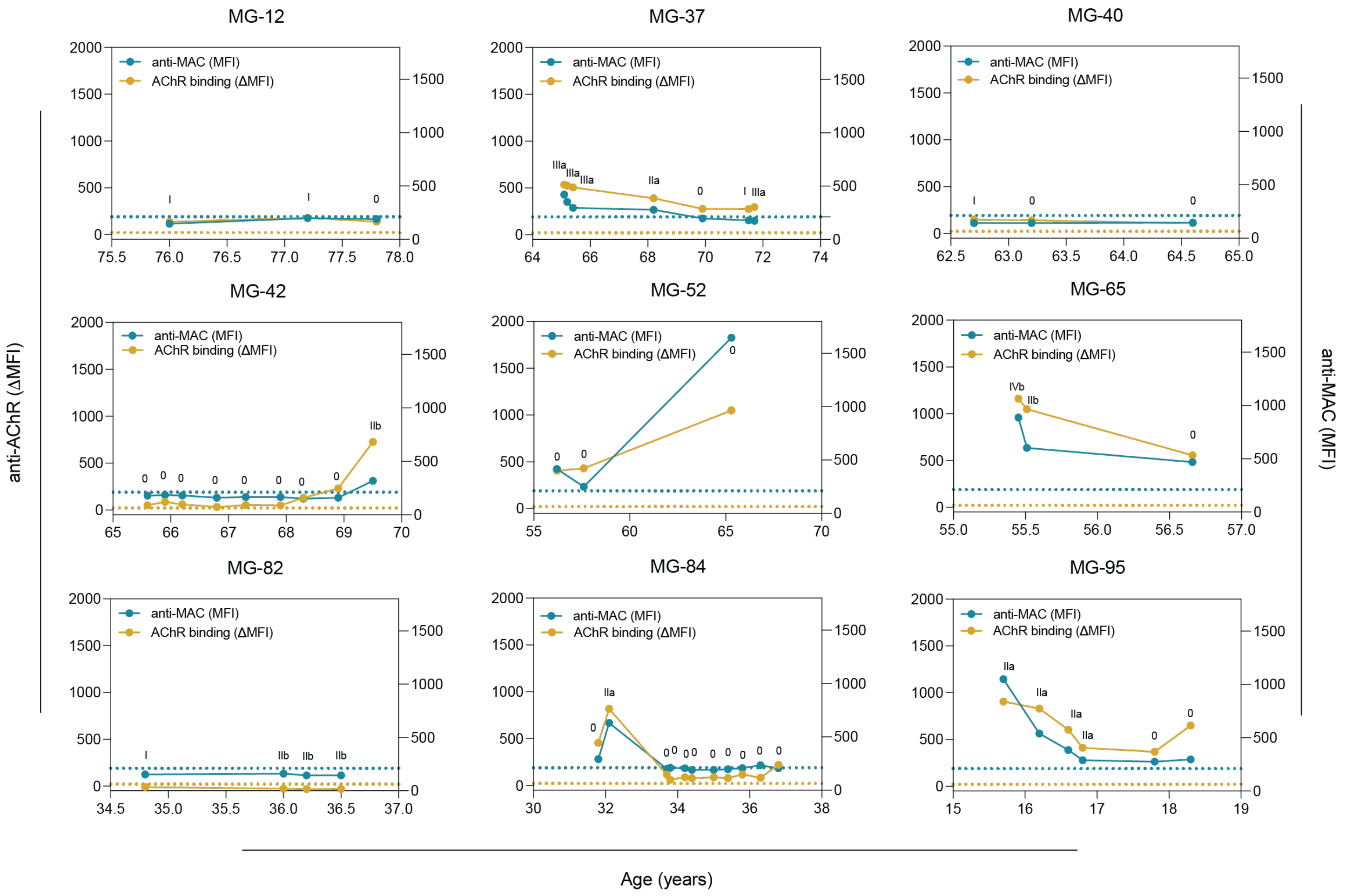

**eFigure 5: Longitudinal changes in AChR autoantibody binding and MAC formation.** Samples, for which at least three serial collections were available (*eTable 1*), were examined side-by-side for both MAC formation and CBA binding, along with MGFA classification corresponding to each timepoint. Each graph shows a single MG patient; the X-axis shows the age of each patient at each collection, the data points show the result for complement CBA (MAC) formation (blue) or CBA binding (golden) MFI. Each data point represents the mean of triplicate experimental conditions. Dotted horizonal lines mark the positive reactivity cut-off for complement (blue) and CBA binding (golden). Cutoffs were calculated using the mean MFI + 4SD of the HD samples (complement and binding CBA 210.9 MFI and 21.9 ΔMFI respectively – see ***Figure 2B***)

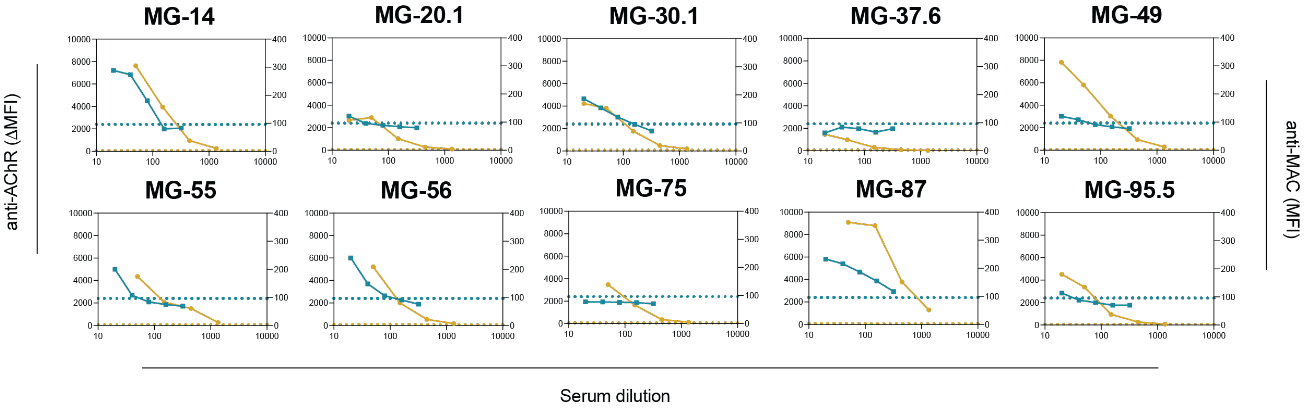

**eFigure 6. Heterogeneous AChR autoantibody-mediated complement formation.** Selected samples (indicated by golden points in ***Figure 2B***) were examined side-by-side for both MAC formation and CBA binding. Each graph shows the data collected from individual serum samples. The X-axis represents the serum dilution of the sample tested for complement CBA (MAC) formation (blue dots) or CBA binding (golden dots). For AChR binding (left y-axis), samples were tested at serum dilutions of 1:20 plus 4 additional 3-fold dilutions (1:50, 1:150, 1:450 and 1:1350). For MAC formation (right y-axis), samples were tested at 2-fold serial dilutions (1:20, 1:40, 1:80, 1:160 and 1:320). Each data point represents the mean of experimental triplicate experimental conditions. Dotted horizonal lines mark the positive reactivity cut-off for complement (blue) and CBA binding (golden). Cutoffs were calculated using the mean MFI + 4STD of the HD samples (complement and CBA 96.01 MFI and 75.35 ΔMFI respectively).

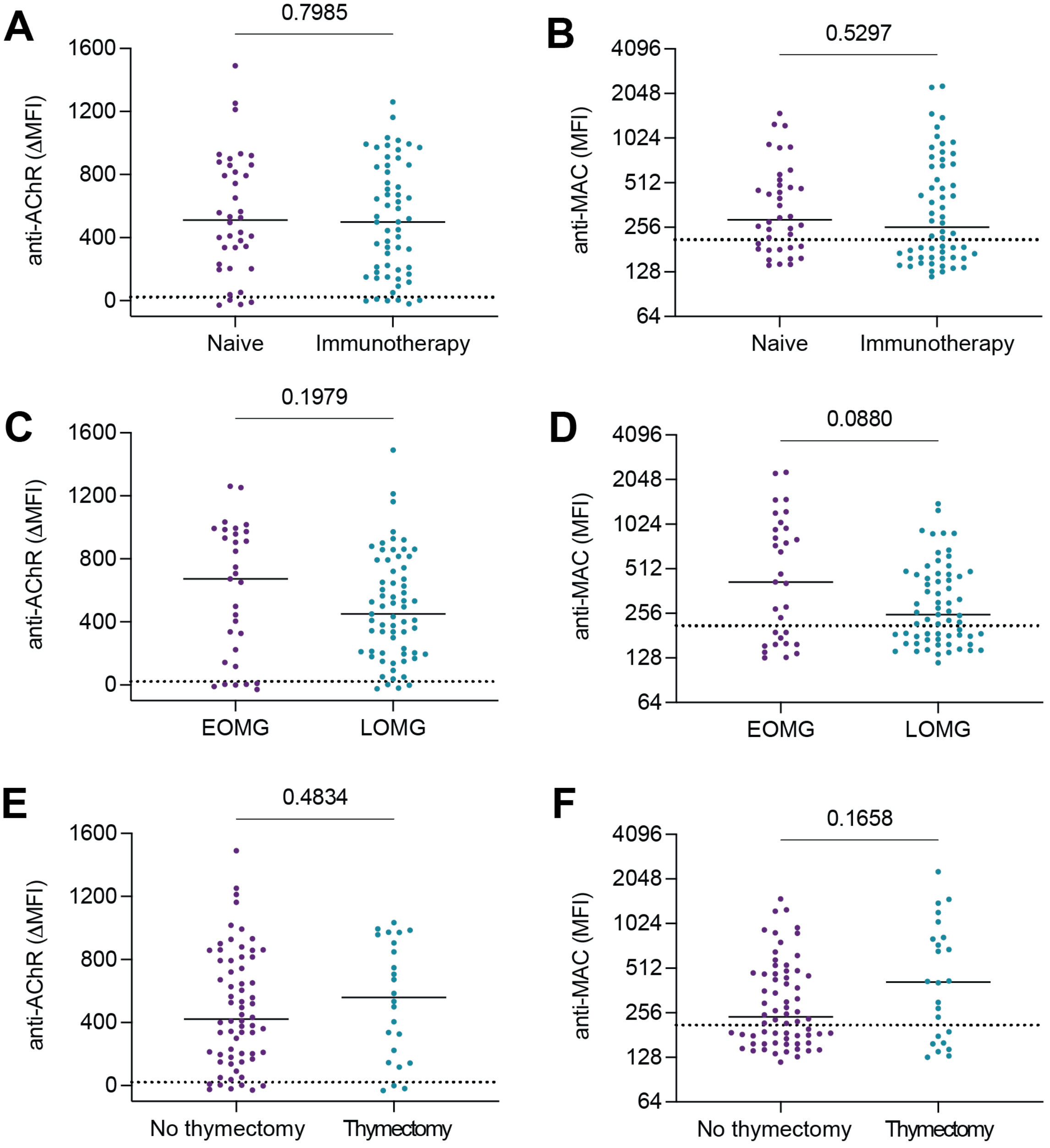

**eFigure 7. CBA binding and MAC formation comparison between MG subgroups.** Cross-sectional samples from various MG subgroups were compared for autoantibody-mediated MAC formation and CBA binding. **A-B.** Immunotherapy naïve patient (n=38) serum samples were compared with patients administered with immunotherapy (n=58), **C-D.** Early on-set MG patient serum (n=31) was compared with late on-set MG patient serum (n=65), **E-F.** MG patients with no thymectomy (n=66) prior to the collection of samples compared with serum samples of patients who had thymectomy (n=24). No significant differences in any of these subgroups were found for both CBA binding and autoantibody mediated MAC-formation. Mann-Whitney U test was used to assess the differences between the groups. Each data point represents the mean of triplicate experimental conditions.

**eTable 1:** Demographics and clinical data of patient and control groups.

| Patient code[1] | Sex | Sample code[2] | Age[3] | EO vs LO[4] | MGFA class[5] | MGC[6] | AChR titer (nmol/L)[7] | anti-MAC (MFI) | AChR binding (ΔMFI) | Immuno-therapy naïve[8] | Current Treatment[9] | Thymec-tomy[10] |
|---|---|---|---|---|---|---|---|---|---|---|---|---|
| 1 | F | 1.0 | 90-100 | LO | IIb | 4 | 1.95 | 184.7 | 300.7 | NO | Imuran, Prednisone, IVIg maintenance, m0estinon | NO |
| 2 | M | 2.0 | 90-100 | LO | IIIb | 17 | 13.7 | 536.0 | 646.3 | NO | None | NO |
| 3 | F | 3.0 | 90-100 | LO | IIa | 12 | 4.75 | 474.7 | 861.7 | YES | Mestinon | NO |
| 4 | M | 4.0 | 90-100 | LO | I | 4 | 9.07 | 439.7 | 793.0 | YES | None | NO |
| 5 | M | 5.0 | 80-89 | LO | IIb | 11 | NA | 347.7 | 721.7 | NO | PLEX | NO |
| 6 | M | 6.0 | 80-89 | LO | I | 3 | 0.3 | 141.7 | -23.7 | YES | None | NO |
| 7 | M | 7.0 | 80-89 | LO | I | 4 | 2.22 | 264.3 | 400.7 | YES | None | NO |
| 8 | M | 8.0 | 80-89 | LO | NA | NA | 1.29 | 296.7 | 408.7 | YES | None | NO |
| 9 | M | 9.0 | 80-89 | LO | V | 42 | 66.5 | 657.3 | 861.3 | NO | Prednisone | NO |
| 10 | M | 10.0 | 80-89 | LO | IIa | 9 | 1.47 | 180.7 | 336.7 | YES | None | NO |
| 11 | M | 11.0 | 80-89 | LO | I | NA | 10.1 | 581.3 | 900.3 | YES | None | NO |
| 12 | M | 12.0 | 70-79 | LO | I | NA | 0.36 | 146.7 | 137.3 | NO | Prednisone | NO |
| | | 12.1 | 70-79 | LO | I | NA | NA | 199.0 | 180.3 | NO | Prednisone | NO |
| | | 12.2 | 70-79 | LO | 0 | NA | 0.46 | 189.7 | 139.3 | NO | Prednisone | NO |
| 13 | M | 13.0 | 70-79 | LO | I | NA | 0.22 | 156.7 | 52.0 | YES | None | NO |
| 14 | F | 14.0 | 70-79 | LO | I | 3 | 14.3 | 1269.0 | 879.7 | YES | Mestinon | NO |
| 15 | F | 15.0 | 70-79 | LO | I | 3 | 4.9 | 217.7 | 337.7 | YES | None | NO |
| 16 | M | 16.0 | 70-79 | LO | IIIa | 13 | 3.54 | 301.0 | 499.3 | NO | Prednisone | YES |
| 17 | M | 17.0 | 70-79 | LO | I | 7 | 0.55 | 187.7 | 150.0 | NO | Prednisone | NO |
| | | 17.1 | 70-79 | LO | I | 2 | 0.55 | 147.3 | 42.7 | NO | Imuran, Prednisone, Mestinon | NO |
| 18 | F | 18.0 | 80-89 | LO | IVb | 18 | 25.3 | 453.7 | 793.0 | YES | None | NO |
| 19 | F | 19.0 | 70-79 | LO | I | 3 | 14.4 | 926.3 | 1489.7 | YES | None | NO |
| 20 | F | 20.0 | 70-79 | LO | IVb | 27 | 7.16 | 428.7 | 1212.7 | YES | Mestinon | NO |
| | | 20.1 | 70-79 | LO | IIb | 5 | 1.23 | 259.0 | 964.0 | NO | Prednisone, IVIg | NO |
| 21 | F | 21.0 | 70-79 | LO | I | 4 | 23.2 | 157.7 | 339.0 | NO | Prednisone, IVIg | NO |
| 22 | M | 22.0 | 70-79 | LO | IIb | 3 | 1.38 | 160.3 | 180.3 | NO | Prednisone, Pembrolizumab, IVIg | NO |
| | | 22.1 | 70-79 | LO | IIa | 7 | 9.51 | 156.7 | 483.3 | NO | IVIg maintenance | NO |
| 23 | M | 23.0 | 70-79 | LO | 0 | 0 | 0 | 118.7 | 3.3 | NO | Prednisone, Azathioprine, | NO |
| 24 | F | 24.0 | 70-79 | LO | NA | 0 | 1.29 | 170.7 | 361.3 | NO | Prednisone, Methotrexate | NO |
| 25 | M | 25.0 | 70-79 | LO | IIIb | NA | 4.85 | 534.3 | 532.7 | YES | Mestinon | NO |
| 26 | M | 26.0 | 70-79 | LO | I | 2 | 1.74 | 217.3 | 449.7 | NO | Prednisone | NO |

| | | | | | | | | | | | | |
|---|---|---|---|---|---|---|---|---|---|---|---|---|
| 27 | M | 27.0 | 70-79 | LO | I | 7 | 7.8 | 491.3 | 651.7 | YES | None | NO |
| 28 | M | 28.0 | 70-79 | LO | IIIb | 11 | 12.8 | 186.7 | 376.0 | NO | Prednisone, IVIg maintenance | NO |
| 29 | M | 29.0 | 70-79 | LO | 0 | 0 | 0.02 | 186.7 | 169.0 | NO | Prednisone | NO |
| 30 | M | 30.0 | 70-79 | LO | I | 1 | 21.3 | 400.7 | 433.3 | YES | Mestinon | NO |
| | | 30.1 | 70-79 | LO | NA | 8 | 92.3 | 842.3 | 692.0 | YES | None | NO |
| 31 | M | 31.0 | 60-69 | LO | I | NA | 4.15 | 359.0 | 410.0 | YES | None | NO |
| | | 31.1 | 70-79 | LO | I | 4 | 24.7 | 695.7 | 940.7 | NO | Imuran, Mestinon | NO |
| 32 | M | 32.0 | 70-79 | LO | 0 | 0 | 5.88 | 251.3 | 557.3 | YES | Imuran, Mestinon | NO |
| 33 | F | 33.0 | 60-69 | LO | I | NA | 0.24 | 229.7 | 230.3 | YES | None | NO |
| 34 | M | 34.0 | 70-79 | LO | IIb | 3 | 1.82 | 232.3 | 627.3 | NO | Prednisone | NO |
| 35 | M | 35.0 | 60-69 | LO | I | NA | 35.2 | 880.3 | 815.7 | YES | Mestinon | NO |
| 36 | F | 36.0 | 70-79 | LO | I | 6 | 1.33 | 248.0 | 527.3 | YES | Mestinon | NO |
| 37 | F | 37.0 | 60-69 | LO | IIIa | NA | 11.7 | 420.0 | 534.7 | NO | Prednisone, Mestinon, IVIg | YES |
| | | 37.1 | 60-69 | LO | IIIa | NA | 12.5 | 352.0 | 524.7 | NO | Rituximab, Prednisone, mestinon | YES |
| | | 37.2 | 60-69 | LO | IIIa | NA | 12.2 | 296.0 | 505.3 | NO | Mestinon, Prednisone | YES |
| | | 37.3 | 60-69 | LO | IIa | 5 | 3.51 | 278.0 | 390.7 | NO | Prednisone | YES |
| | | 37.4 | 60-69 | LO | 0 | 0 | 2.19 | 197.7 | 277.7 | NO | Prednisone, Mestinon | YES |
| | | 37.5 | 70-79 | LO | I | 3 | 5.46 | 178.7 | 275.0 | NO | Prednisone | YES |
| | | 37.6 | 70-79 | LO | IIIa | 20 | 64.7 | 175.0 | 296.3 | NO | Prednisone, Mestinon | YES |
| 38 | M | 38.0 | 60-69 | LO | IIIb | NA | 10.3 | 302.7 | 380.3 | YES | None | NO |
| 39 | M | 39.0 | 60-69 | LO | 0 | NA | 0.92 | 147.3 | 211.3 | NO | IVIg | NO |
| | | 39.1 | 60-69 | LO | IVa | 28 | 13.2 | 1436.0 | 1088.3 | NO | Prednisone, PLEX | NO |
| 40 | M | 40.0 | 60-69 | LO | I | NA | 0.95 | 141.7 | 149.3 | NO | Imuran, Prednisone | NO |
| | | 40.1 | 60-69 | LO | 0 | NA | 0.47 | 141.3 | 141.0 | NO | Rituximab, Imuran, Prednisone | NO |
| | | 40.2 | 60-69 | LO | 0 | NA | 0.4 | 146.3 | 109.9 | NO | None | NO |
| 41 | M | 41.0 | 60-69 | LO | IIb | 3 | 5.02 | 377.3 | 858.0 | NO | None | NO |
| 42 | M | 42.0 | 60-69 | LO | 0 | 0 | 0.99 | 179.0 | 51.3 | NO | Rituximab, Prednisone | NO |
| | | 42.1 | 60-69 | LO | 0 | NA | NA | 185.0 | 88.3 | NO | Rituximab, Prednisone | NO |
| | | 42.2 | 60-69 | LO | 0 | 0 | 0.77 | 180.3 | 60.7 | NO | Rituximab, Prednisone | NO |
| | | 42.3 | 60-69 | LO | 0 | 0 | 0.46 | 159.0 | 32.0 | NO | Prednisone | NO |
| | | 42.4 | 60-69 | LO | 0 | 0 | 0.2 | 165.7 | 53.3 | NO | None | NO |
| | | 42.5 | 60-69 | LO | 0 | 0 | 0.51 | 165.0 | 49.3 | NO | None | NO |
| | | 42.6 | 60-69 | LO | 0 | NA | 1.03 | 150.0 | 130.0 | NO | None | NO |
| | | 42.7 | 60-69 | LO | 0 | 0 | 1.32 | 160.0 | 231.3 | NO | None | NO |
| | | 42.8 | 60-69 | LO | IIb | 8 | 9.94 | 317.0 | 725.7 | NO | None | NO |

| | | | | | | | | | | | | |
|---|---|---|---|---|---|---|---|---|---|---|---|---|
| 43 | M | 43.0 | 60-69 | LO | I | 7 | 14.3 | 182.7 | 495.0 | YES | Mestinon | NO |
| 44 | M | 44.0 | 60-69 | LO | I | I | 1.47 | 179.3 | 344.7 | YES | None | NO |
| 45 | M | 45.0 | 60-69 | LO | V | NA | NA | 318.3 | 604.7 | NO | IVIg, IV solumedrol, PLEX | NO |
| 46 | F | 46.0 | 60-69 | LO | I | 4 | 2.72 | 198.3 | 203.7 | YES | Imuran, Mestinon | NO |
| 47 | M | 47.0 | 60-69 | LO | I | 7 | 7.59 | 466.3 | 565.0 | YES | None | NO |
| 48 | M | 48.0 | 60-69 | LO | I | 3 | 49.2 | 1408.3 | 972.3 | NO | Imuran, Prednisone | YES |
| 49 | M | 49.0 | 60-69 | LO | III | 16 | 2.54 | 260.0 | 744.0 | YES | None | NO |
| 50 | M | 50.0 | 60-69 | LO | IIb | 3 | 8.08 | 160.0 | 214.0 | NO | Prednisone, Mestinon | NO |
| 51 | M | 51.0 | 60-69 | LO | IVb | 17 | 9.33 | 468.3 | 816.0 | NO | Prednisone | NO |
| 52 | F | 52.0 | 50-59 | EO | 0 | NA | 3.03 | 416.0 | 405.3 | NO | Prednisone | YES |
| | | 52.1 | 50-59 | EO | 0 | 0 | 2.84 | 249.3 | 431.0 | NO | None | YES |
| | | 52.2 | 60-69 | EO | 0 | 0 | 10.3 | 1647.3 | 1049.7 | NO | None | YES |
| 53 | M | 53.0 | 60-69 | LO | I | 5 | 0 | 139.3 | -19.3 | NO | Prednisone, Cellcept, IVIg maintenance | NO |
| 54 | F | 54.0 | 60-69 | LO | IIb | 6 | 147 | 623.7 | 858.7 | YES | None | NO |
| 55 | M | 55.0 | 60-69 | LO | 0 | 4 | 11.4 | 684.0 | 585.3 | NO | IVIg maintenance | YES |
| 56 | M | 56.0 | 60-69 | LO | IVb | 28 | 88 | 277.7 | 927.7 | YES | Mestinon | NO |
| 57 | F | 57.0 | 60-69 | LO | IIb | 13 | 1.17 | 145.3 | -1.3 | NO | Prednisone, Mestinon, IVIg | NO |
| 58 | F | 58.0 | 50-59 | EO | IIb | NA | NA | 1217.7 | 748.2 | NO | Imuran, Prednisone | YES |
| 59 | M | 59.0 | 50-59 | LO | I | 7 | 3.17 | 186.3 | 203.0 | YES | None | NO |
| | | 59.1 | 50-59 | LO | 0 | 0 | 8.2 | 170.7 | 289.3 | NO | Prednisone, IVIg maintenance | NO |
| 60 | F | 60.0 | 50-59 | LO | I | 4 | NA | 170.0 | 196.0 | NO | Prednisone, Mestinon, Cellcept | NA |
| 61 | M | 61.0 | 50-59 | LO | 0 | 0 | 0 | 158.0 | 92.3 | NO | Prednisone, Mestinon | NO |
| | M | 61.1 | 50-59 | LO | I | 8 | NA | 189.0 | 373.7 | NO | None | NO |
| 62 | F | 62.0 | 50-59 | EO | NA | NA | 44.8 | 2254.3 | 1261.3 | NO | Mestinon | NA |
| | | 62.1 | 50-59 | EO | IIIb | NA | 3.31 | 279.0 | 489.3 | NO | Imuran, Prednisone, IVIg | NA |
| 63 | F | 63.0 | 50-59 | EO | IIa | NA | 123 | 1500.3 | 1252.7 | YES | Mestinon | NO |
| 64 | M | 64.0 | 50-59 | LO | IIIb | 0 | 13.5 | 889.7 | 920.7 | YES | None | NO |
| 65 | M | 65.0 | 50-59 | LO | IVb | 23 | 35.7 | 886.7 | 1162.7 | NO | Prednisone, Mestinon | NO |
| | | 65.1 | 50-59 | LO | IIb | 10 | 15.2 | 602.7 | 1048.7 | NO | Prednisone, Mestinon, IVIg | NO |
| | | 65.2 | 50-59 | LO | 0 | 0 | 15.8 | 468.3 | 557.2 | NO | Prednisone, Mestinon | NO |
| 66 | M | 66.0 | 50-59 | LO | IIIb | 10 | 9.7 | 223.0 | 519.3 | NO | Prednisone, Mestinon | NO |

| | | | | | | | | | | | | |
|---|---|---|---|---|---|---|---|---|---|---|---|---|
| 67 | F | 67.0 | 50-59 | LO | IIa | 2 | 4.9 | 490.7 | 671.3 | NO | Prednisone, Mestinon, Cellcept | NO |
| 68 | M | 68.0 | 50-59 | LO | I | 3 | 100 | 143.7 | 197.0 | YES | None | NO |
| 69 | F | 69.0 | 40-49 | EO | I | 2 | 2.7 | 764.7 | 1017.0 | NO | Methotrexate | NO |
| 70 | F | 70.0 | 50-59 | EO | IIb | 12 | 16.6 | 733.0 | 848.0 | NO | Cellcept, Mestinon, IVIg maintenance | YES |
| 71 | M | 71.0 | 40-49 | EO | IIa | 9 | 0 | 157.7 | -28.3 | YES | None | NO |
| 72 | F | 72.0 | 40-49 | EO | IIIb | 17 | 11.5 | 824.7 | 986.3 | NO | Prednisone, Cellcept, Mestinon | YES |
| | | 72.1 | 40-49 | EO | 0 | 0 | 7.29 | 625.0 | 713.3 | NO | Prednisone, Cellcept, IVIg maintenance | YES |
| 73 | F | 73.0 | 40-49 | EO | 0 | NA | 5.05 | 190.3 | 327.3 | NO | Imuran | YES |
| | | 73.1 | 40-49 | EO | IIb | 2 | 16.8 | 183.0 | 270.7 | NO | None | YES |
| 74 | F | 74.0 | 40-49 | EO | IIIa | 19 | NA | 665.3 | 957.3 | NO | None | YES |
| 75 | F | 75.0 | 40-49 | EO | IIB | 10 | 11.3 | 174.3 | 652.0 | NO | Prednisone, Imuran, IVIg maintenance, Mestinon | NA |
| 76 | F | 76.0 | 30-39 | EO | 0 | 0 | 3.73 | 408.3 | 707.0 | NO | Imuran | YES |
| 77 | F | 77.0 | 30-39 | EO | 0 | NA | 0.27 | 128.3 | 0.0 | NO | IVIg maintenance | YES |
| 78 | M | 78.0 | 60-69 | LO | I | 1 | 0.46 | 144.3 | 37.7 | YES | None | NO |
| 79 | F | 79.0 | 30-39 | EO | IIIb | 13 | 39 | 1493.3 | 994.7 | NO | Prednisone, IVIg | YES |
| | | 79.1 | 40-49 | EO | IIIb | 19 | 22.2 | 1095.0 | 990.3 | NO | Cellcept | YES |
| | | 79.2 | 40-49 | EO | IIIb | 7 | 22.8 | 1196.3 | 1115.7 | NO | Prednisone, Mestinon, Cellcept, IVIg | YES |
| 80 | F | 80.0 | 30-39 | EO | I | NA | NA | 129.3 | 10.0 | NO | Prednisone | NO |
| | | 80.1 | 30-39 | EO | I | 5 | 0 | 160.0 | 0.0 | NO | Prednisone | NO |
| 81 | F | 81.0 | 30-39 | EO | 0 | 0 | 20 | 137.0 | 3.6 | NO | Mestinon, IVIg | NO |
| | | 81.1 | 30-39 | EO | IIa | 7 | 8.14 | 131.0 | -18.0 | NO | Mestinon, IVIg | YES |
| 82 | F | 82.0 | 30-39 | EO | I | 5 | 0.41 | 154.3 | -9.4 | YES | Mestinon | NO |
| | | 82.1 | 30-39 | EO | IIb | 11 | 0.22 | 161.0 | -26.7 | NO | Prednisone, Mestinon | NO |
| | | 82.2 | 30-39 | EO | IIb | 8 | 0.25 | 145.0 | -32.0 | NO | Prednisone, IVIg maintenance | YES |
| | | 82.3 | 30-39 | EO | IIb | 6 | 0.25 | 144.0 | -31.0 | NO | Prednisone, IVIg maintenance | YES |
| 83 | F | 83.0 | 30-39 | EO | IIIb | NA | 0.61 | 472.7 | 498.3 | NO | Mestinon | NA |
| 84 | F | 84.0 | 30-39 | EO | 0 | NA | 4.15 | 282.7 | 444.0 | NO | Prednisone | NO |
| | | 84.1 | 30-39 | EO | IIa | 3 | 9.19 | 667.7 | 763.3 | NO | Prednisone | NO |
| | | 84.2 | 30-39 | EO | 0 | 0 | 0.82 | 177.7 | 145.7 | NO | Rituximab, Prednisone | YES |
| | | 84.3 | 30-39 | EO | 0 | 0 | 0.48 | 189.3 | 97.7 | NO | Rituximab, Prednisone | YES |
| | | 84.4 | 30-39 | EO | 0 | NA | 0.64 | 185.7 | 121.8 | NO | Rituximab, Prednisone | YES |
| | | 84.5 | 30-39 | EO | 0 | 0 | 0.72 | 167.0 | 112.7 | NO | Prednisone | YES |

| | | | | | | | | | | | | |
|---|---|---|---|---|---|---|---|---|---|---|---|---|
| | | 84.6 | 30-39 | EO | 0 | 0 | 0.73 | 168.3 | 122.0 | NO | Prednisone | YES |
| | | 84.7 | 30-39 | EO | 0 | 0 | 0.97 | 174.0 | 114.3 | NO | Prednisone | YES |
| | | 84.8 | 30-39 | EO | 0 | 0 | 2.37 | 186.0 | 148.0 | NO | Prednisone | YES |
| | | 84.9 | 30-39 | EO | 0 | 0 | 3.28 | 217.3 | 118.3 | NO | None | YES |
| | | 84.10 | 30-39 | EO | 0 | 0 | 2.79 | 184.7 | 238.3 | NO | None | YES |
| 85 | M | 85.0 | 30-39 | EO | I | 0 | 0 | 190.3 | 4.0 | YES | Mestinon | NO |
| | | 85.1 | 30-39 | EO | I | 1 | 0 | 151.3 | -3.0 | NO | Prednisone, Mestinon | NO |
| 86 | M | 86.0 | 30-39 | EO | 0 | 2 | 0.01 | 158.0 | 142.7 | NO | Imuran | YES |
| 87 | F | 87.0 | 30-39 | EO | IIa | 5 | 43.1 | 806.0 | 1034.7 | NO | Prednisone, Mestinon | YES |
| 88 | M | 88.0 | 30-39 | EO | IIIa | 15 | 27.4 | 1241.0 | 931.7 | YES | None | NO |
| | | 88.1 | 30-39 | EO | IIb | 8 | 45.5 | 1263.0 | 906.7 | NO | Prednisone | NA |
| 89 | F | 89.0 | 30-39 | EO | IIa | NA | 89.8 | 2290.0 | 973.5 | NO | Mestinon | YES |
| 90 | F | 90.0 | 30-39 | EO | IIa | 2 | NA | 942.0 | 912.3 | NO | Mestinon, IVIg maintenance | NA |
| 91 | F | 91.0 | 30-39 | EO | IVb | 31 | 2.47 | 239.0 | 673.3 | NO | Rituximab, Mestinon, IVIg maintenance | YES |
| | | 91.1 | 30-39 | EO | IIIb | 28 | 0.83 | 187.3 | 558.3 | NO | Rituximab, IVIg maintenance | YES |
| 92 | F | 92.0 | 20-29 | EO | 0 | NA | 6.61 | 274.3 | 336.7 | NO | None | YES |
| 93 | M | 93.0 | 20-29 | EO | IVb | 20 | 10.1 | 960.0 | 993.0 | NO | Prednisone, Azathioprine, | NO |
| 94 | F | 94.0 | 20-29 | EO | IIIb | 12 | 0.68 | 160.0 | 223.4 | NO | Mestinon | YES |
| | | 94.1 | 20-29 | EO | I | 1 | 0.49 | 155.3 | 288.0 | NO | Prednisone, IVIg | YES |
| 95 | M | 95.0 | 10-19 | EO | IIa | 4 | 12 | 1049.3 | 905.3 | NO | Prednisone, IVIg maintenance | YES |
| | | 95.1 | 10-19 | EO | IIa | 0 | 12 | 539.3 | 830.7 | NO | Rituximab, Prednisone, IVIg maintenance | YES |
| | | 95.2 | 10-19 | EO | IIa | 4 | 6.66 | 384.7 | 604.7 | NO | Rituximab, Prednisone, IVIg maintenance | YES |
| | | 95.3 | 10-19 | EO | IIa | 1 | 8.89 | 289.0 | 411.7 | NO | Rituximab, Prednisone, IVIg maintenance | YES |
| | | 95.4 | 10-19 | EO | 0 | 0 | 5.75 | 275.7 | 369.7 | NO | Mestinon | YES |
| | | 95.5 | 10-19 | EO | 0 | 0 | 10 | 296.0 | 649.0 | NO | None | YES |
| 96 | M | 96.0 | 10-19 | EO | 0 | 0 | 0.1 | 140.0 | 117.3 | NO | Prednisone, Mestinon | YES |

**Healthy donors (HD)**

| **HD code[1]** | **Sex** | **Sample code[2]** | **Age[3]** |
|---|---|---|---|
| 1 | F | 1 | 20-29 |
| 2 | F | 2 | 30-39 |
| 3 | M | 3 | 30-39 |

| | | | | | |
|---|---|---|---|---|---|
| 4 | M | 4 | 30-39 | | |
| 5 | M | 5 | 40-49 | | |
| 6 | M | 6 | 20-29 | | |
| 7 | F | 7 | 20-29 | | |
| 8 | M | 8 | 20-29 | | |
| 9 | F | 9 | 20-29 | | |
| 10 | F | 10 | 60-69 | | |
| 11 | M | 11 | 60-69 | | |
| 12 | M | 12 | 30-39 | | |
| 13 | F | 13 | 30-39 | | |
| 14 | M | 14 | 30-39 | | |
| 15 | M | 15 | 20-29 | | |
| 16 | M | 16 | 20-29 | | |
| 17 | F | 17 | 50-59 | | |
| 18 | M | 18 | 40-49 | | |
| 19 | M | 19 | 30-39 | | |
| 20 | M | 20 | 30-39 | | |
| 21 | M | 21 | 30-39 | | |
| 22 | M | 22 | 20-29 | | |
| 23 | F | 23 | 30-39 | | |
| 24 | M | 24 | 20-29 | | |
| 25 | F | 25 | 20-29 | | |
| 26 | M | 26 | 20-29 | | |
| 27 | F | 27 | 20-29 | | |
| 28 | F | 28 | 20-29 | | |
| 29 | M | 29 | 20-29 | | |
| | | 29.1 | 20-29 | | |
| 30 | F | 30 | 20-29 | | |
| | | 30.1 | 20-29 | | |
| 31 | F | 31 | 50-59 | | |
| 32 | M | 32 | 30-39 | | |
| 33 | F | 33 | 20-29 | | |
| 34 | M | 34 | 50-59 | | |
| 35 | F | 35 | 20-29 | | |
| 36 | M | 36 | 30-39 | | |
| **Neuromyelitis optica spectrum disorder (NMOSD)** | | | | | |
| **Patient code[1]** | **Sex** | **Sample code[2]** | **Age[3]** | **Autoantibody status** | **Immunotherapy naïve[8]** |
| 1 | F | 1 | 60-69 | AQP4+ | YES |

Abbreviations: M = Male; F = Female; NA = Not available, EO = Early-onset MG; LO = Late-onset MG

[1, 2] Each patient code consists of either one or multiple longitudinal serum samples. Longitudinal samples are listed in chronological order.

[3] Patient age range (years) at time of sample collection.

[4] Early onset vs late onset MG was determined by age of diagnosis: early-onset (<50 years); late-onset MG (≥50 years).

[5] The MGFA Clinical Classification.

[6] The MG composite (MGC) score.
[7] Serum sample AChR titer (nmol/L) as measured by RIA.
[8] Immunotherapy includes cellcept, prednisone, azathioprine, methotrexate, IV solumedrol, IV immunoglobulin, rituximab and PLEX.
[9] Treatment at time of sample collection. Rituximab treatment is reported if it is within 6 months of sample collection. IVIg and PLEX treatment reported if administered within 12 weeks of sample collection. IVIg maintenance or PLEX maintenance is reported if treatment administration is recurring.
[10] Thymectomy status at time of sample collection.

**eTable 2: CRISPR gRNA sequences for the generation of CD46/55/59 KO HEK293T**

| Gene | NCBI gene ID | Primer Name | gRNA sequence (5'-3') |
|---|---|---|---|
| hCD46 | 4179 | Anti | TCTAGACAGGGGGTCACCGT |
| | | Sense | AAAGTATGAAGACCATGTAG |
| hCD55 | 1604 | Sense | (G)AAATGACTCCCACCCGAACA |
| | | Sense | (G)AGAAAATAGTGTCAATCAGA |
| hCD59 | 966 | Anti | GTGGCAGTGCCTGTAAACCA |
| | | Sense | GCTGAGCTCAAAGGACACGT |
